# Reduction in the 2020 Life Expectancy in Brazil after COVID-19

**DOI:** 10.1101/2021.04.06.21255013

**Authors:** Marcia C Castro, Susie Gurzenda, Cassio M Turra, Sun Kim, Theresa Andrasfay, Noreen Goldman

## Abstract

Brazil has the second-largest number of COVID-19 deaths worldwide. We use data on reported deaths to measure and compare the death toll across states from a demographic perspective. We estimate a decline in 2020 life expectancy at birth of 1.94 years, resulting in a mortality level not seen since 2013. The reduction in life expectancy at age 65 was 1.58 years, setting Brazil back to 2009 levels. The decline was larger for males, widening by 2.3% and 5.4% the female-male gap in life expectancy at birth and at age 65, respectively. Among states, Amazonas lost 59.6% of the improvements in life expectancy at birth since 2000. With 2021 COVID-19 deaths at about 43% of the total 2020 figures (as of mid-March) the demographic effect is likely to be even higher this year.

## Main Text

As of March 23, COVID-19 has officially claimed more than 2.7 million lives worldwide, 48% of which are in the Americas. The actual death toll is likely to be higher due to deficient surveillance, limited testing that prevented proper diagnosis, issues with compliance to protocols on reporting a suspected COVID-19 death, and location of the death (e.g., at home) *(1, 2)*. Together, the United States and Brazil account for 31% of the world’s death toll and 64% of the Americas’. In both countries, the pandemic response in 2020 was disparate regionally, with lamentable national coordination *(3, 4)*, resulting in a high and unequal mortality burden *(5)*. Currently, Brazil faces the worst moment of the pandemic, with increasing cases, deaths, and hospitalizations. The number of COVID-19 deaths reported on March 23, 2021, accounted for 33% worldwide.

The consequences of that death toll can be measured in terms of life expectancy at birth, which indicates the average number of years a newborn would be expected to live if born in a specific year and subjected to the prevailing mortality rates in that year throughout life. Changes in life expectancy at birth can reflect differences in expected longevity between two periods, such as prior to versus during a pandemic. For example, the 1918 influenza pandemic was estimated to have reduced life expectancy at birth in the United States by 7 to 12 years *(6)*. Mortality due to COVID-19 is estimated to have reduced the United States life expectancy at birth by 1.13 years, setting it back to values observed in 2003. That decline shows large disparities by race/ethnicity: a reduction of 3.05, 2.10, and 0.68 years in life expectancy at birth among the Latino, Black, and White populations, respectively *(7)*. Considering the large COVID-19 death toll in Brazil, with marked regional inequalities, the goal of this study is to quantify the loss in years of life expectancy at birth and at age 65 due to the pandemic. Estimates are detailed by sex and by states, and we measure and compare changes in the regional and female-male gaps in life expectancy.

Before COVID-19, life expectancy at birth in Brazil still lagged behind many countries in Asia, Europe, and the Americas. In Latin America, at least four countries experienced secular mortality declines at earlier dates: Argentina, Uruguay, Costa Rica, and Cuba *(8)*. Between 1945 and 2020, life expectancy at birth in Brazil increased from 45.5 to 76.7 *(9)*, an average of almost five months per calendar year. This relatively fast pace characterized many countries that had been lagging their peers in mortality improvement and subsequently benefitted from their forerunners’ progress *(8, 10)*.

Based on the total number of COVID-19 deaths reported in Brazil in 2020, we estimated a reduction of 1.94 years in the life expectancy at birth (**Fig. 1A and Table S1**), with a larger drop for males (1.98 years) compared to females (1.82 years). This result is 72% higher than the estimated decline for the United States *(7)*. The Federal District, Brazil’s capital, had an estimated decrease in the life expectancy at birth of 3.68 years, the highest absolute decline among all states. Overall, declines were higher in the North region, led by Amapá (3.62 years), Roraima (3.43 years), and Amazonas (3.28 years) (**Fig. 1B**). We also estimated changes in life expectancy at age 65 (**Fig. S1 and Table S2**), given the higher risk of dying from COVID-19 at older ages *(5)*. The estimated decline for Brazil was 1.58 years for both sexes, 1.46 for females, and 1.64 for males. Across states, the largest declines were estimated for the Federal District (3.08 years), Amapá (2.98 years), Amazonas (2.92 years), and Roraima (2.74 years). Male life expectancy at both birth and age 65 had a larger percentage decline than females in all states, reflecting the pattern of higher risk of dying from COVID-19 among men (**Table S2 and Fig. S2**) *(5)*. In the majority of states, the absolute decline was also larger among males (**Table S2**).

**Fig. 1.**
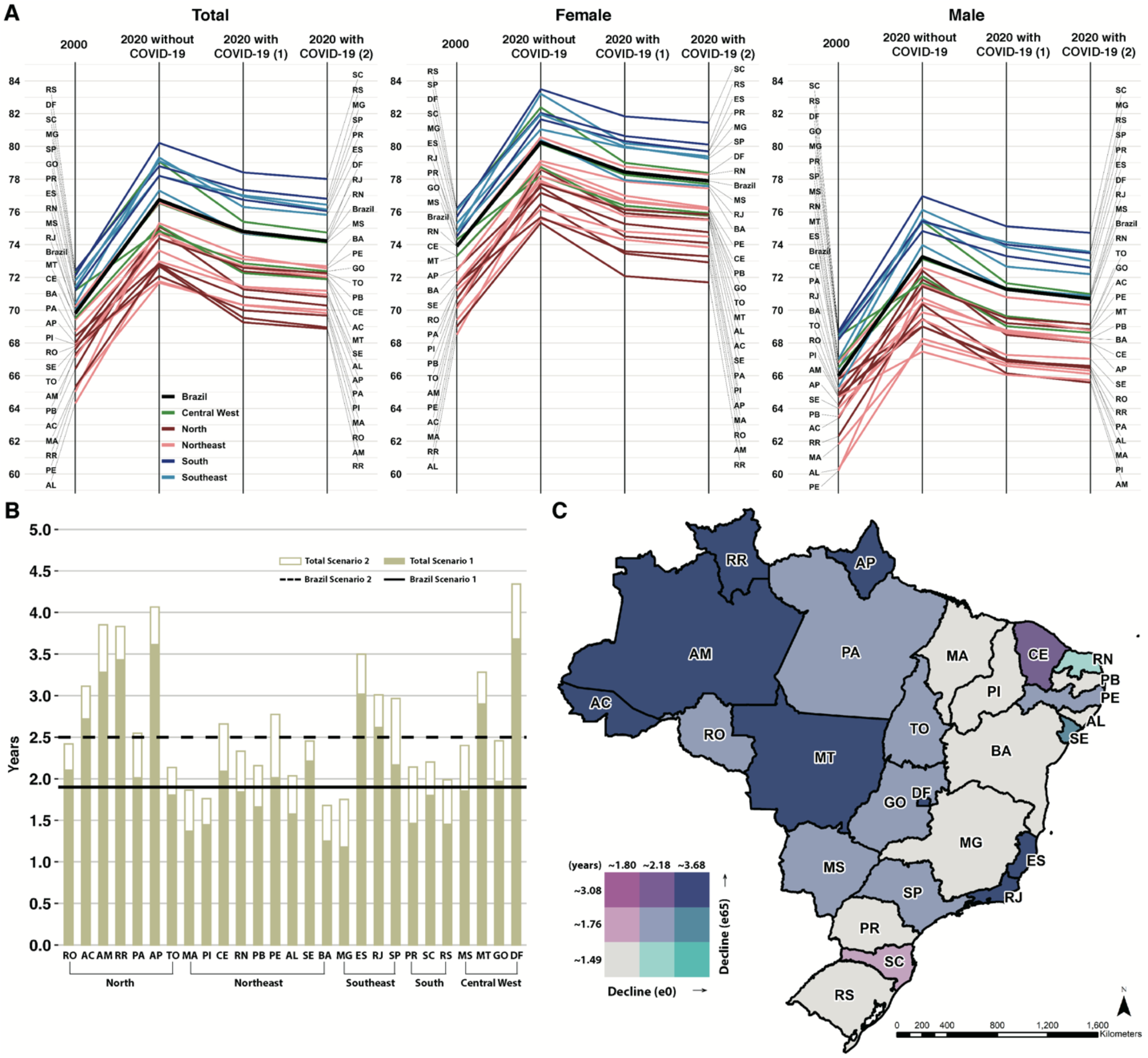
Changes in life expectancy at birth by state and sex. **(A)** Life expectancy at birth in 2000, 2020 without COVID-19, and considering COVID-19 mortality under two alternative scenarios. Estimates by state and sex. Scenario (1) considers all COVID-19 deaths reported in 2020. Scenario (2) assumes that, during the COVID-19 pandemic, 90% of the severe acute respiratory infections (SARI) deaths without a confirmed diagnosis were COVID-19 deaths (see supplementary materials). States are colored according to major regions. State acronyms by region, North: AC=Acre, AP=Amapá, AM=Amazonas, PA=Pará, RO=Rondônia, RR=Roraima, and TO=Tocantins; Northeast: AL=Alagoas, BA=Bahia, CE=Ceará, MA=Maranhão, PB=Paraíba, PE=Pernambuco, PI=Piauí, RN=Rio Grande do Norte, and SE=Sergipe; Center-West: DF=Distrito Federal, GO=Goiás, MT=Mato Grosso, and MS=Mato Grosso do Sul; Southeast: ES=Espírito Santo, MG=Minas Gerais, RJ=Rio de Janeiro, and SP=São Paulo; South: PR=Paraná, RS=Rio Grande do Sul, and SC=Santa Catarina. **(B)** Decline (in years) in life expectancy at birth due to COVID-19 for both sexes (total) under two alternative scenarios. **(C)** Bivariate choropleth map of the decline (in years) in life expectancy at birth and at age 65.

On average, larger declines in life expectancy were observed in the North, and smaller in the South and Northeast regions (**Fig. 1C**). States in the North and Northeast regions have the worst indicators of income inequality, poverty, access to infrastructure, and availability of physicians and hospital beds *(11)*. In the Northeast, however, the estimated decreases in life expectancy are smaller (**Fig. 1**). Governors of the states in that region imposed the most rigorous measures of physical distancing, in direct opposition to recommendations from the president *(12)*.

We considered an alternative scenario of COVID-19 mortality to account for probable misdiagnosis in reported severe acute respiratory infections (SARI). Specifically, we assumed that, during the COVID-19 pandemic, 90% of the SARI deaths without a confirmed diagnosis were COVID-19 deaths. Under that scenario, the decline in life expectancy at birth estimated for Brazil would be 2.52 years for both sexes, 2.37 for females, and 2.56 for males (**Fig 1 and Table S3**). Concerning life expectancy at age 65, we estimated a reduction of 2.04 years for Brazil, with the largest decline of 3.75 years for males in Amapá (**Fig. S1 and Table S4**).

To understand how these changes affected the gap in life expectancy between males and females, we compared the estimated gap in 2020 without COVID-19 with the estimated gap considering COVID-19 mortality. Our results point to an overall widening of the male-female gap in life expectancy at birth of 0.16 years in Brazil (0.19 under the alternative scenario). In Amapá, the male-female gap is estimated to increase by 0.91 years (**Fig. 2A and Table S5**). Larger male-female gaps were estimated for life expectancy at age 65 (**Fig. 2B and Table S5**), an increase of 0.18 years in Brazil, 1.01 years in Amapá, and 0.45 years in Amazonas.

**Fig. 2.**
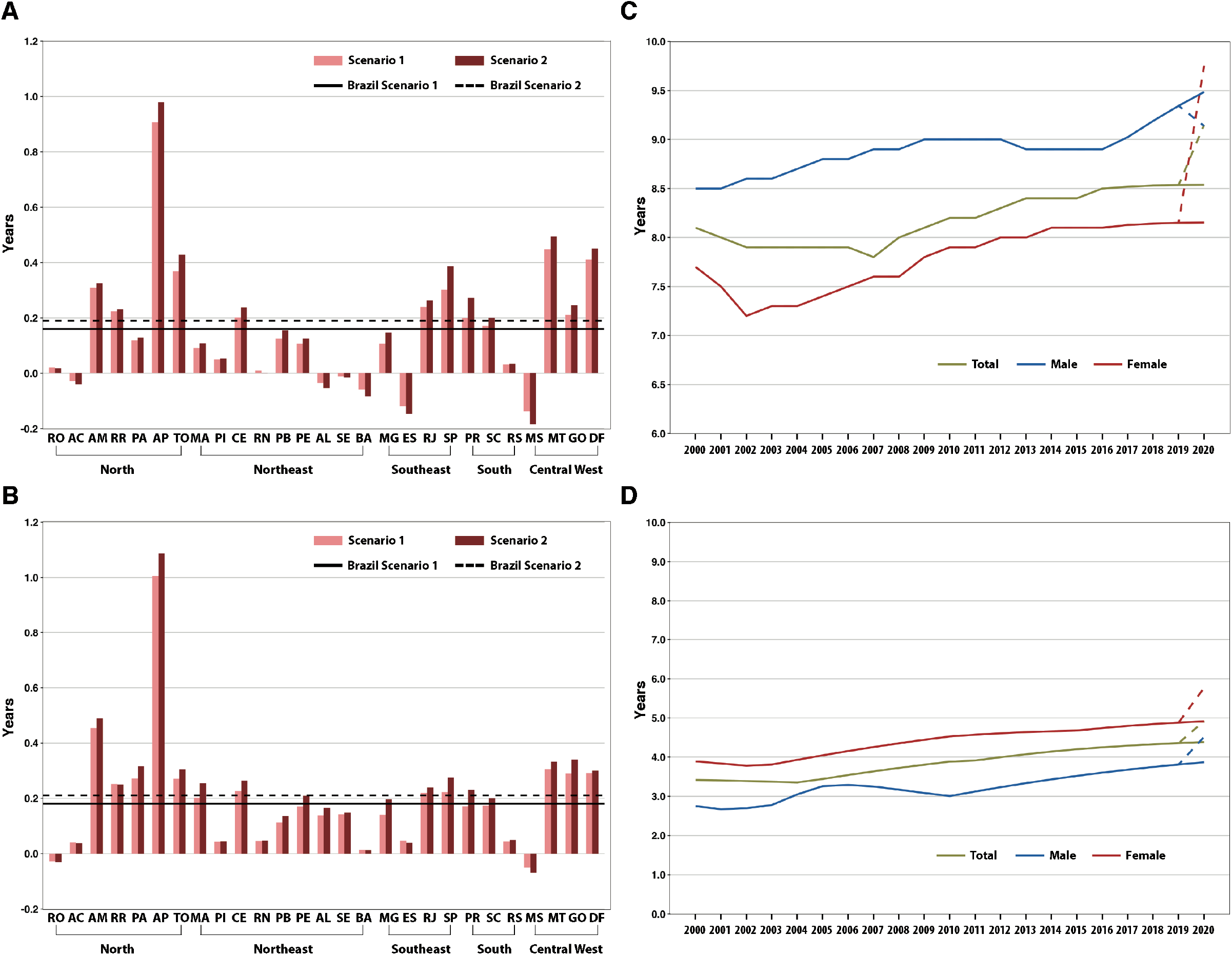
Changes in gaps of life expectancy at birth and age 65 by state and sex. Change (in years) in the 2020 female-male gap in life expectancy at birth **(A)** and age 65 **(B)** under two alternative scenarios of COVID-19 mortality. Lines represent changes for Brazil. Bars, organized by major regions, show changes for the states. State acronyms by region, North: AC=Acre, AP=Amapá, AM=Amazonas, PA=Pará, RO=Rondônia, RR=Roraima, and TO=Tocantins; Northeast: AL=Alagoas, BA=Bahia, CE=Ceará, MA=Maranhão, PB=Paraíba, PE=Pernambuco, PI=Piauí, RN=Rio Grande do Norte, and SE=Sergipe; Center-West: DF=Distrito Federal, GO=Goiás, MT=Mato Grosso, and MS=Mato Grosso do Sul; Southeast: ES=Espírito Santo, MG=Minas Gerai, RJ=Rio de Janeiro, and SP=São Paulo; South: PR=Paraná, RS=Rio Grande do Sul, and SC=Santa Catarina. Regional gap (in years) in life expectancy at birth **(C)** and age 65 **(D)**, calculated as the difference between the highest and smallest life expectancy among states. Dotted lines are the estimated gap given COVID-19 deaths (see supplementary materials).

We also calculated regional inequalities in life expectancy, or the difference between the lowest and the highest state life expectancy, to assess changes due to COVID-19. The difference in life expectancy at birth increased from 8.54 to 9.15 years, and at age 65 from 4.38 to 4.92 years (**Fig. 2C and 2D, and Table S6**). The regional inequity for females had the largest estimated increase: 1.59 years for life expectancy at birth, and 0.86 years for life expectancy at age 65. Among males, the difference also increased at age 65 (0.65 years). However, it declined by 0.41 years when measured at birth. In this case, Piauí (the state with the lowest male life expectancy at birth before COVID-19, located in the Northeast region) experienced a relatively small decline in life expectancy (**Table S6**). These results suggest that COVID-19 mortality increased regional inequalities, reflecting the pandemic’s disproportional burden among vulnerable groups *(5)* but also spatial patterns of COVID-19 spread *(13)*.

We quantified the effect of COVID-19 on reversing the progress in mortality reduction since 2000. Of the gains achieved in life expectancy at birth in Brazil over two decades, more than a quarter was lost due to COVID-19 (**Table S7 and Fig. 3A**), and 36% under the alternative scenario (**Table S7 and Fig. S3A**). Among the states, Amazonas lost approximately 60% for both sexes, and 70% among males (**Table S7**). In 11 states (including the entire Southeast region), this loss was higher among females (**Fig. 3A**). A much higher loss was estimated for life expectancy at age 65, almost half for both sexes in Brazil (**Table S8**). Among males, only five states had losses below 50%, and four lost more than 100%, including Amazonas (**Fig. 3B**).

**Fig. 3.**
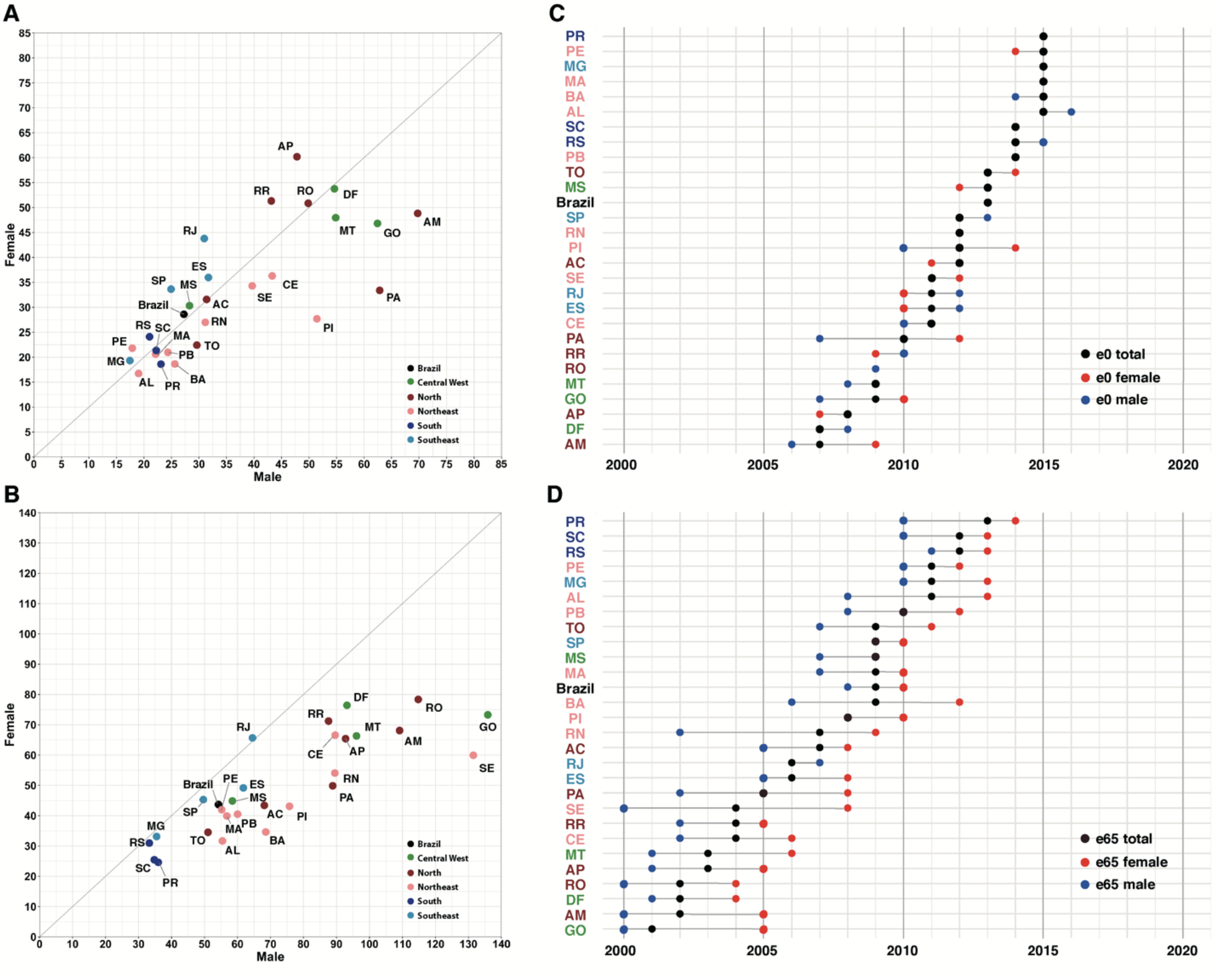
Loss and setback in life expectancy at birth and at age 65 by state and sex. Percentage loss due to COVID-19 mortality relative to increases in female and male life expectancy at birth **(A)** and age 65 **(B)** between 2000 and 2020, by state. States are colored according to major regions. State acronyms by region, North: AC=Acre, AP=Amapá, AM=Amazonas, PA=Pará, RO=Rondônia, RR=Roraima, and TO=Tocantins; Northeast: AL=Alagoas, BA=Bahia, CE=Ceará, MA=Maranhão, PB=Paraíba, PE=Pernambuco, PI=Piauí, RN=Rio Grande do Norte, and SE=Sergipe; Center-West: DF=Distrito Federal, GO=Goiás, MT=Mato Grosso, and MS=Mato Grosso do Sul; Southeast: ES=Espírito Santo, MG=Minas Gerais, RJ=Rio de Janeiro, and SP=São Paulo; South: PR=Paraná, RS=Rio Grande do Sul, and SC=Santa Catarina. Setback in life expectancy at birth **(C)** and age 65 **(D)** due to COVID-19 mortality, by sex and state (see supplementary materials). State acronyms are colored according to major regions. In some states the setback is similar among total, males, or females (**Table S9**).

As a result, estimated life expectancy at birth in Brazil in the presence of COVID-19 reflects levels observed in 2013 (2011 under the alternative scenario) (**Table S9, and Fig. 3C and S3**). Among the states, the largest setback was estimated for Amazonas (levels of 2007 for both sexes, 2009 for females, and 2006 for males). The estimated setback for life expectancy at age 65 is overwhelming, equalling values last observed as far back as 2001 in most states and prior to 2000 in a few (including Amazonas and the Federal District) (**Fig. 3D**).

Our results quantify the COVID-19 death toll in Brazil from a demographic perspective. We show that the life expectancy in the presence of COVID-19 is equivalent to levels observed in Brazil between five and over 20 years ago, depending on the state. While these findings are disturbing, we argue that they are almost certainly underestimated. First, our alternative scenario corrects only for misdiagnosis among SARI deaths. It is possible that COVID-19 was the main cause of some deaths that happened at home, and others for which the cause of death was not properly reported due to lack of testing. Second, mortality due to all other causes may have changed as well, for two reasons. One is that those who survived COVID-19 could have become more vulnerable to other conditions, in the short-run, due to previous co-morbidities. A second factor is that postponement of medical tests and procedures, avoidance in seeking emergency care, interruption of primary care, and inferior quality of care for treatment of conditions other than COVID-19 due to hospital overcrowding could have prompted fatal complications. Indeed, not all estimated excess mortality in Brazil in 2020 can be accounted for by the number of reported COVID-19 deaths *(1)*.

When intense shocks like a pandemic or war occur, life expectancy drops but often rebounds quickly. This was the case with the 1918 influenza pandemic in the United States, when life expectancy in 1919 was higher than in 1917, likely due in part to selective mortality of individuals with tuberculosis *(14)*. We argue that, in the case of COVID-19 in Brazil, the rebound will not happen immediately. We offer five reasons as to why. First, Brazil is currently going through the worst moment of the pandemic, with a record high and increasing number of deaths and a very slow immunization rollout. The hospital system is facing an unprecedented collapse. Twenty-five of the 27 capital cities have intensive care unit (ICU) occupancy above 80% (19 of those above 90%) *(15)*. As of March 14, 2021, the number of COVID-19 deaths in Brazil is 42.7% of those reported in 2020. In Amazonas, COVID-19 deaths in 2021 already surpassed the 2020 figures (**Fig. 4A and Table S10**). Manaus, Amazonas’ capital, observed a very high attack rate after the first wave of the pandemic in 2020 *(16)*. Recently, it witnessed one of the most tragic scenarios of overwhelmed hospital capacity, running out of not only hospital beds but also oxygen *(17)*. A new variant of concern (P.1), which emerged in the city in November 2020, is estimated to be 1.4-2.2 times more transmissible than other lineages, and may evade protected immunity from a previous infection *(18)*. As of March 15, 2021, this variant has been detected in 16 states *(19)*. Therefore, the death toll in 2021 may well exceed that in 2020.

**Fig. 4.**
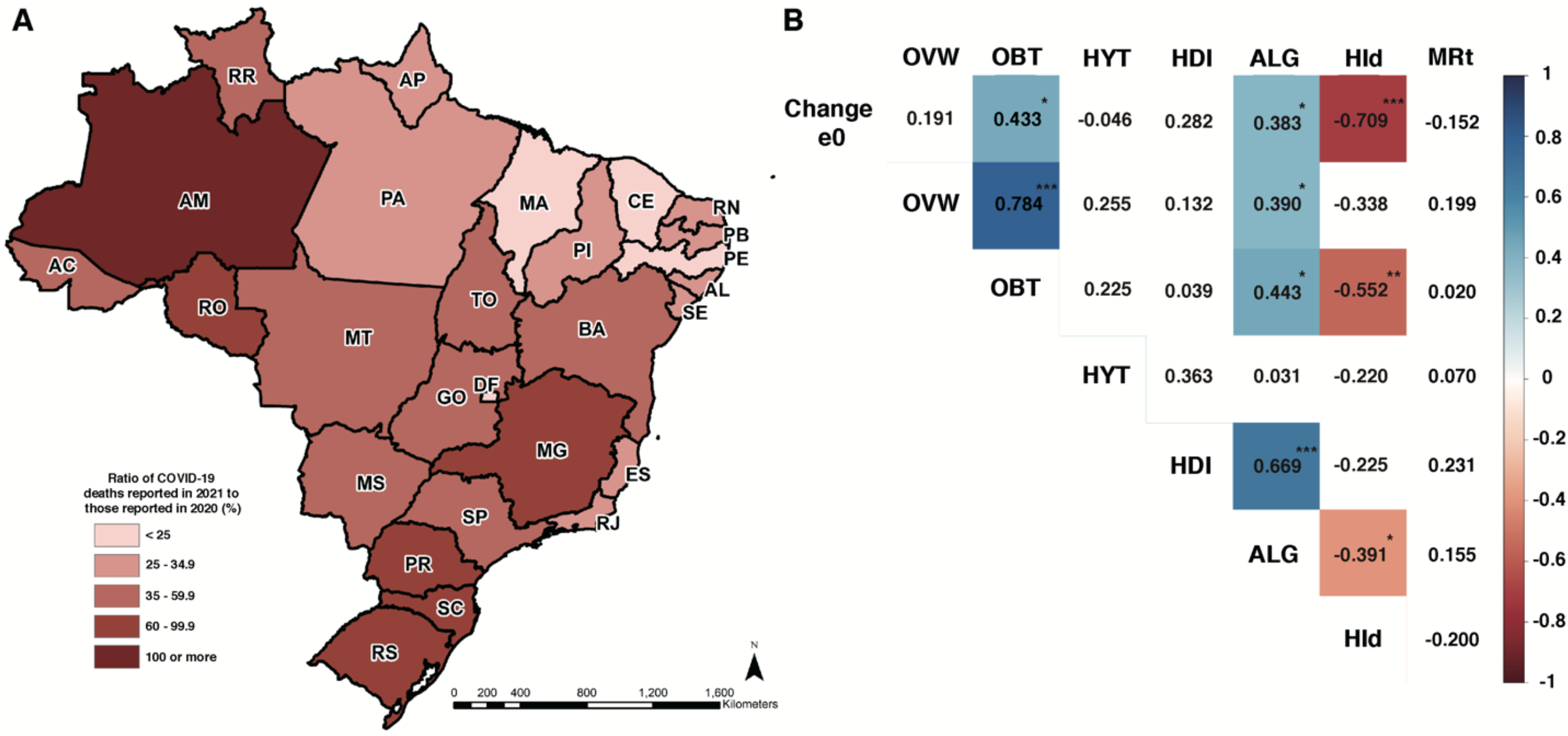
Indicators of the COVID-19 death toll. **(A)** Ratio of COVID-19 deaths reported in 2021 (as of March 14) to those reported in 2020 (%). Numbers above 100 indicate more deaths in 2021 than 2020. State acronyms by region, North: AC=Acre, AP=Amapá, AM=Amazonas, PA=Pará, RO=Rondônia, RR=Roraima, and TO=Tocantins; Northeast: AL=Alagoas, BA=Bahia, CE=Ceará, MA=Maranhão, PB=Paraíba, PE=Pernambuco, PI=Piauí, RN=Rio Grande do Norte, and SE=Sergipe; Center-West: DF=Distrito Federal, GO=Goiás, MT=Mato Grosso, and MS=Mato Grosso do Sul; Southeast: ES=Espírito Santo, MG=Minas Gerais, RJ=Rio de Janeiro, and SP=São Paulo; South: PR=Paraná, RS=Rio Grande do Sul, and SC=Santa Catarina. **(B)** Correlation matrix (Pearson). Variables (all state-level): estimated change in the life expectancy at birth in 2020 due to COVID-19 mortality (Change e0), percentage of adults who are overweight in the capital city (OVW), percentage of adults who are obese in the capital city (OBT), percentage of adults with hypertension in the capital city (HYT), human development index (HDI), political alignment between the governor and the president, as indicated by the governor’s support during the 2018 presidential elections (ALG), median Hoover Index of COVID-19 deaths (HId), and maximum effective reproductive number in 2020 (MRt). Cells in shades of red or blue are statistically significant: * <0.05, ** <0.01, and *** <0.001 (see supplementary materials).

Second, COVID-19 disrupted primary care services in Brazil *(20)*. This compromised screening for cancer, with a reduction of about 35% in new diagnoses *(21)*. Child immunization was reduced, particularly among impoverished children in the North region *(22)*. Disruption in treatment and diagnosis of tuberculosis and HIV may increase mortality over the next five years *(23)*. Overall health conditions of individuals with diabetes worsened in 2020 due to reduced physical activity, postponement of medical appointments, and interruption in regular drug treatment *(24)*. These are some examples of deteriorating health conditions that not only will generate a higher demand for health care services but may also affect future mortality patterns.

Third, reports of long-term consequences of COVID-19 among survivors continue to emerge *(25, 26)*. These include fatigue and neurological, pulmonary, and cardiovascular complications. At present, there is no concrete evidence that COVID-19 sequelae may shorten individual life span, but close monitoring to assess deterioration of health conditions and interaction with other pre-existing co-morbidities is needed.

Fourth, an economic crisis hit Brazil in 2015. Since then, poverty and inequality have increased *(27)*. COVID-19 exacerbated this scenario, exposing the most vulnerable to food insecurity *(28)*. From April to December 2020, an emergency basic income program mitigated the challenges imposed by the pandemic (e.g., unemployment). However, the basic income benefit ended in December 2020. The first three months of 2021 have been the most critical since COVID-19 emerged in Brazil, with increasing and record-high cases, deaths, and hospitalizations. Many cities are imposing lockdowns. Yet, no financial support is currently provided to a growing population living in poverty. A more limited version of the basic income program may be re-established in April 2021.

Fifth, reductions in the health budget and changes in the health-financing model are likely to affect health outcomes *(29)*. They may reduce access to and coverage of primary care, and increase infant mortality and avoidable deaths. Ultimately, inequality may become worse, exacerbating an already distressing scenario due to COVID-19.

Previously we described the spatiotemporal patterns of the spread of COVID-19 cases and deaths in Brazil. We showed that a largely unmitigated pandemic, in a context of local inequalities, resulted in a high and unequal mortality burden *(13)*. The demographic magnitude of the COVID-19 death toll is not homogenous across states and is associated with the pattern of spread *(13)*. We find that the faster the speed at which COVID-19 spread across municipalities (a lower locational Hoover Index), the larger the changes in life expectancy *(13)* (**Fig. 4B and Table S11**). We also show a positive correlation between the prevalence of obesity in the state capital cities and declines in life expectancy. To assess pandemic response, we used a simple proxy of political alignment: whether a state governor supported the current president in the last election. Our results show a positive correlation between political alignment and losses in life expectancy at birth.

In summary, the death toll of COVID-19 in Brazil in 2020 has been catastrophic. State-level gains in longevity achieved over years or even decades were reversed by the pandemic. The lack of a coordinated, prompt, and equitable response informed by science, as well as the promotion of disinformation, have been the hallmark of the current administration *(20)*. Brazil does not lack a universal health care system, or a network of community health agents to target vulnerable communities, or minimum data and a capable cadre of researchers tirelessly advancing knowledge and informing policy. It lacks leadership commitment to save lives. After more than 195,000 lives were reported to have been lost in 2020 to COVID-19, no policy changes have appeared in 2021. As many countries speed up vaccination coverage, and witness declines in cases and deaths, Brazil is moving in the opposite direction. Without a change in coordination of pandemic response, immediate lockdowns to contain the current surge, and a rapid increase in administering vaccines, Brazil will soon become a serious threat to national and global health security *(30)*. The consequences, sadly and unacceptably, will continue to be measured in human lives lost, and the future demographic consequences may be even worse than those reported here.

## Data Availability

The data and code required to reproduce the results in this article can be found on Zenodo (available upon acceptance).

## Author contributions

Conceptualization – MCC and NG. Methodology – MCC, NG, and TA. Software – TA, SG, and SK. Formal analysis – MCC, SG. Interpretation – MCC, NG, and CMT. Data curation – MCC and SG. Drafting – MCC and CMT. Review and editing – all authors. Visualization – MCC and SK. Funding – MCC.

## Data and materials availability

The data and code required to reproduce the results in this article can be found on Zenodo (available upon acceptance). Since data were de-identified, this study did not involve human subjects. This work is licensed under a Creative Commons Attribution 4.0 International (CC BY 4.0) license, which permits unrestricted use, distribution, and reproduction in any medium, provided the original work is properly cited. To view a copy of this license, visit https://creativecommons.org/licenses/by/4.0/. This license does not apply to figures/photos/artwork or other content included in the article that is credited to a third party; obtain authorization from the rights holder before using such material.

## Materials and Methods

### Data

We used several data sources to estimate the changes in life expectancy due to COVID-19 by state, sex, and age in Brazil. We obtained abridged life tables and mid-year population projections for 2020 by state, age, and sex from the Brazilian Institute of Geography and Statistics *(Instituto Brasileiro de Geografia e Estatística*, IBGE, in Portuguese) (https://www.ibge.gov.br/en/statistics/social/population/18176-population-projection.html). To estimate the COVID-19 scenario, we used confirmed COVID-19 deaths in 2020 as reported by Brasil.io, which compiles epidemiological bulletins from the 27 State Health Departments. These deaths, aggregated at the state level, are publicly available by date on their website (https://brasil.io/covid19/). We abstracted death data on February 13, 2021, for all states and the Federal District, as reported through December 31, 2020 (N = 195,072 confirmed COVID-19 deaths). These data are not available by age and sex.

To obtain age and sex structure, we used de-identified publicly available data on severe acute respiratory illness (SARI) hospitalizations from the Influenza Epidemiological Surveillance Information System *(Sistema de Informação de Vigilância Epidemiológica da Gripe*, SIVEP-Gripe, in Portuguese). The data are frequently updated and made publicly available by the Ministry of Health (https://opendatasus.saude.gov.br/nl/dataset). We used the February 8, 2021 version. This dataset provides the age and sex structure of hospitalized COVID-19 deaths. In-hospital deaths represent 91% of all COVID-19 deaths reported in 2020. SIVEP-Gripe also reports whether a diagnosis is “unspecified” which can be interpreted as not identified as COVID-19, influenza, or another severe acute respiratory illness.

### Estimating COVID-19 Deaths

Our analysis consists of three scenarios: the base-case and two COVID-19 scenarios. The base-case estimates the 2020 life expectancies in the absence of COVID-19. The first COVID-19 scenario (Scenario 1) includes all COVID-19 deaths reported in 2020. The second (Scenario 2) attempts to adjust for underreporting of COVID-19 deaths among SARI deaths, as detailed next.

The deaths in the base-case scenario are deaths that were projected to occur in 2020 in the absence of COVID-19 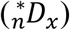. We calculated those deaths by applying projected age group- and sex-specific mortality rates for 2020 from each state to the corresponding projected population. We used the 2020 mortality rates in the abridged life tables published by IBGE. We also used IBGE’s 2020 mid-year population projections by state, sex, and five-year age group.

Scenario 1 comprises all deaths in the base-case plus COVID-19 confirmed deaths 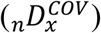 in 2020, as reported by Brasil.io. In the absence of age and sex details for reported COVID-19 deaths, we apply the age and sex structure of confirmed COVID-19 in-hospital deaths in 2020 by state. We estimate the total deaths in 2020, inclusive of COVID-19 deaths, to be:

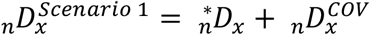

where 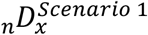 are all deaths that are estimated to occur in 2020 in Scenario 1, in the age range x to x + n, 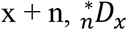 are deaths in the absence of COVID-19, and 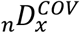 are deaths associated with confirmed COVID-19.

Scenario 2 attempts to correct for some of the underreporting in COVID-19 deaths *(1)*. Specifically, we considered that, during a pandemic, unspecified deaths due to severe acute respiratory infections (SARI) should be about 10% of total SARI deaths. We assumed that in 2020, 90% of the unspecified SARI deaths in each state were unidentified COVID-19 deaths. Therefore, the corrected number of in-hospital deaths associated with COVID-19 was calculated as the sum of confirmed COVID-19 in-hospital deaths and 90% of the unspecified in-hospital SARI deaths in each state. Next, we estimated the state-specific proportion of total reported COVID-19 deaths that were hospitalized. The corrected number of COVID-19 deaths (regardless of hospitalization status) was calculated as the product of the corrected number of hospitalized COVID-19 deaths and the inverse of the proportion of confirmed COVID-19 deaths that were hospitalized.

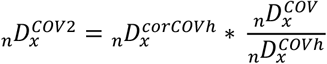

where 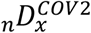 is the estimated number of COVID-19 deaths accounting for underreporting, 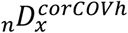 is the corrected number of hospitalized COVID-19 deaths (accounting for 90% of unspecified SARI deaths by state in 2020), 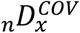 is the number of confirmed COVID-19 deaths in 2020, and 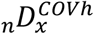 is the number of in-hospital deaths associated with confirmed COVID-19 in 2020. Thus, the total number of age-specific deaths in scenario 2 can be calculated as:

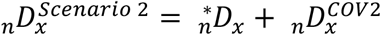

Data curation and analysis were conducted in Stata version 15.1 (Stata Corp., College Station, TX, USA) and R version 4.0.0 (R core team, 2020).

### Changes in Life Expectancy

We constructed life tables using conventional demographic methods that considered COVID-19 deaths by state, sex, and age group. In the base-case scenario, we consider expected deaths in 2020 in the absence of COVID-19 and use the original abridged IBGE life tables for 2020 by state and sex, aggregated to match this analysis’s age intervals. Previous work explains this age group aggregation process in detail *(7, 31)*. We treat these base-case life tables as the cause-deleted life tables (also known as associated single decrement life tables). These are hypothetical life tables assuming that a single cause of death was eliminated *(32)*. In this case, COVID-19 is the eliminated cause. Using the base-case life tables as cause-deleted life tables, we then construct all-cause life tables for causes of death inclusive of confirmed COVID-19 deaths (Scenario 1) and the corrected number of COVID-19 deaths, accounting for underreporting (Scenario 2). Prior studies have used this approach to estimate COVID-19 life tables *(7, 31)*.

For each scenario, we calculated _*n*_*R*_*x*_, the age- and sex-specific ratio of deaths in the absence of COVID-19 to deaths with COVID-19. This ratio is similar to that described by Chiang’s method: the ratio of deaths from a single cause to deaths from all causes *(33)*. However, here we replaced the variable for deaths from a single cause with the variable deaths from all but one cause, so the ratio represents deaths from all but one cause to deaths from all causes.

We then applied standard life table relationships to complete the life tables in the presence of COVID-19 *(32)*. We calculated the probability of surviving from age x to age x + n (_*n*_*p*_*x*_), the probability of dying between ages x and x + n (_*n*_*q*_*x*_), and the average person-years lived in the interval by those dying in the interval (_*n*_*a*_*x*_) in the presence of COVID-19. We used Chiang’s method, which, as Andrasfay and Goldman previously noted, assumes that the force of decrement from cause *i* (or in this case, all causes except *i)* is proportional to the force of decrement from all other causes *(7, 33)*.

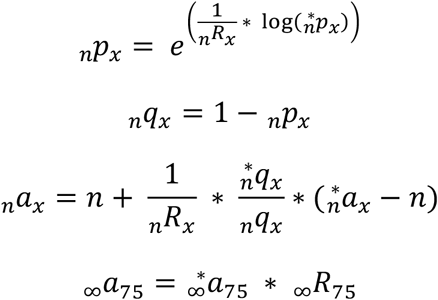

These 2020 life expectancy calculations were performed for each sex and state combination for Scenarios 1 and 2 (input data and results available on Zenodo - available upon acceptance).

The change in life expectancy due to COVID-19 deaths is estimated by subtracting the life expectancy considering deaths associated with COVID-19 at a specific age from the life expectancy estimated without considering COVID-19. We tabulated these changes at birth and at age 65 for Scenarios 1 and 2 (**Tables S1 to S4**). We calculated changes in the female-male gap in life expectancy at birth and at age 65 (**Table S5**). Also, we estimated the difference between the highest and lowest state life expectancies (**Table S6**). We further quantified the loss due to COVID-19 considering the increases in life expectancy at birth and at age 65 (by sex and state) over the past two decades (2000 to 2020) (**Tables S7 and S8**). Lastly, we measured the setback in life expectancy due to COVID-19 mortality by identifying the year when the life expectancies estimated in Scenarios 1 and 2 were last observed in Brazil and each state (**Table S9**).

All demographic analyses were done in Microsoft Excel version 16.44 with additional data cleaning and processing in Stata version 15.1 (Stata Corp., College Station, TX, USA) and R version 4.0.0 (R core team, 2020). Data visualizations were created in R.

### Comparison of COVID-19 Death Rate in 2020 and 2021

To compare the effect of the pandemic in 2020 to the current state in 2021 (as of March 14), we calculated the death rate in both years, accounting for person-time of exposure. The death rate for a particular state and year is calculated as the number of deaths divided by the number of person-years lived at risk. Person-years is calculated as the mid-year population times the number of years of exposure to a certain risk (in this case COVID-19). For 2020, we measured exposure time as the number of days remaining in 2020 after the first case in each state. Because at the time of writing there are active cases in all states, the exposure time in 2021 was measured from January 1 through March 14. Brazil and state crude death rates are shown in **Table S10**.

### Correlations

We assessed the strength of associations between eight variables using the Pearson correlation coefficient (**Fig 4B**). The variables are:

1. Estimated change in the life expectancy at birth in 2020 due to COVID-19 mortality considering Scenario 1 (Change e0) – **Table S1**
2. Percentage of overweight adults in each state capital city in 2019 (OVW) – **Table S11**
3. Percentage of obese adults in each state capital city in 2019 (OBT) – **Table S11**
4. Percentage of individuals with hypertension in each state capital city in 2019 (HYT) – **Table S11**
5. Human development index (HDI) in 2017 – **Table S11**
6. Political alignment between the governor and the president, as indicated by the governor’s support during the 2018 presidential elections (ALG) – **Table S11**
7. Median locational Hoover Index of COVID-19 deaths (HId) in 2020 – **Table S11**
8. Maximum effective reproductive number in 2020 (MRt) – **Table S11**

Percentage of adults reporting being overweight (OVW, Body Mass Index (BMI) ≥ 25kg/m), obese (OBT, Body Mass Index (BMI) ≥ 30kg/m), and hypertensive (HYT) were obtained from an annual phone survey conducted by the Ministry of Health in Brazil, called VIGITEL *(34)*. The survey assesses risk factors for chronic diseases and is representative of all capital cities in Brazil. Here we used the data for each capital city as a proxy for the state. HDI was obtained from Brazil Atlas *(35)*. It captures three dimensions of development: longevity, education, and income. We used the latest HDI published for states in Brazil (for the year 2017). ALG was obtained from Pereira et al *(11)*. The locational Hoover Index is a measure of spatial imbalance between two variables. We previously calculated the weekly index for COVID-19 deaths by states in Brazil *(13)*. Here we use the median value. Lastly, the maximum estimated value of the effective reproduction number *(R*_*t*_) in 2020 (MRt) was extracted from *Observatório Covid-19 BR* (https://covid19br.github.io/), a multidisciplinary group of researchers that have been working together to produce varied indicators and to disseminate knowledge on the pandemic.

All calculations and visualizations were done in R (R core team, 2020).

**Fig. S1.**
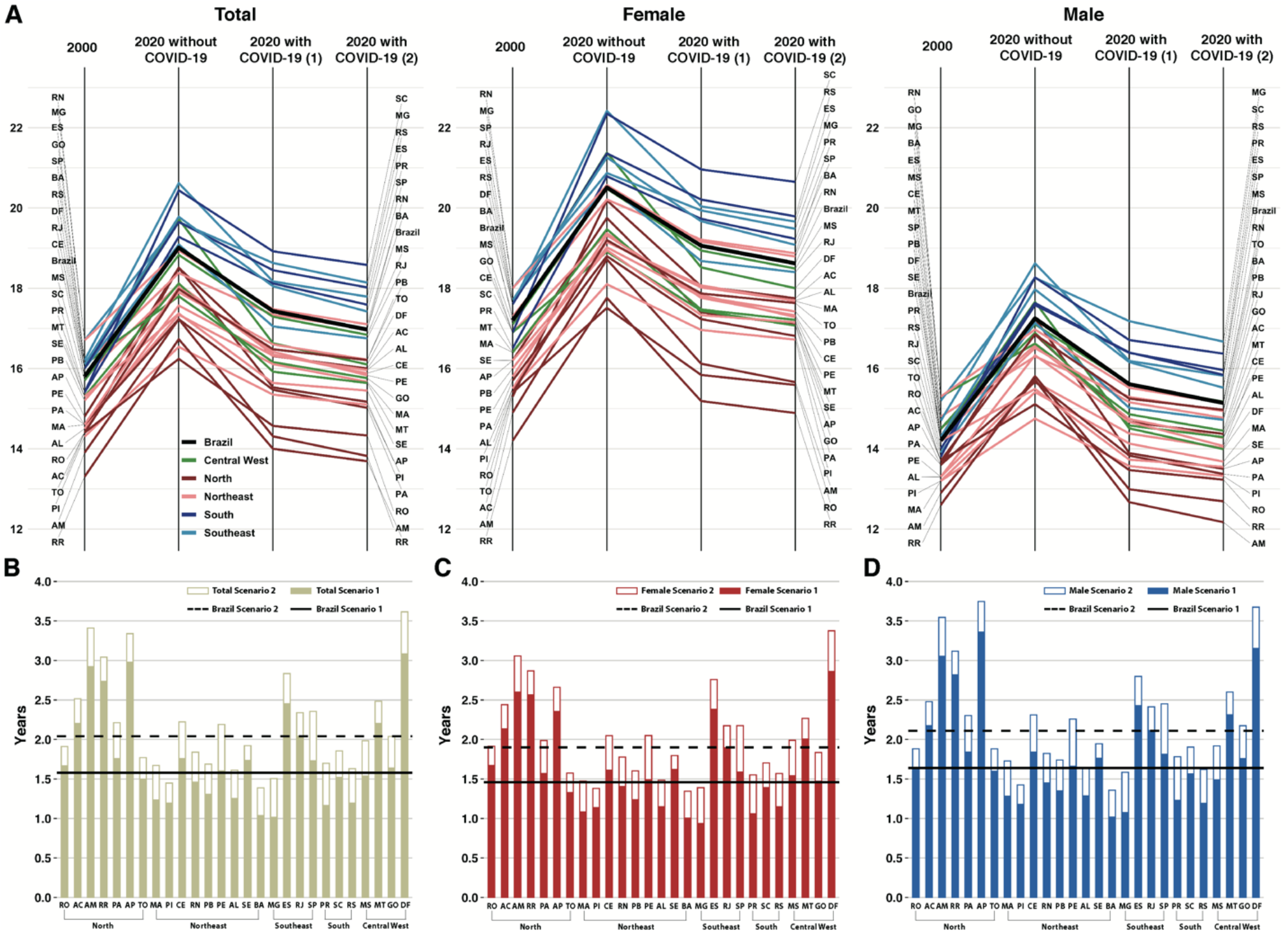
Changes in life expectancy at age 65 by state and sex. **(A)** Life expectancy at age 65 in 2000, 2020 without COVID-19, and considering COVID-19 mortality under two alternative scenarios. Estimates by state and sex. Scenario (1) considers all COVID-19 deaths reported in 2020. Scenario (2) assumes that, under a pandemic, 90% of the unspecified severe acute respiratory infections (SARI) deaths in each state were unidentified COVID-19 deaths (see **Materials and Methods**). States are colored according to major regions. State acronyms by region, North: AC=Acre, AP=Amapá, AM=Amazonas, PA=Pará, RO=Rondônia, RR=Roraima, and TO=Tocantins; Northeast: AL=Alagoas, BA=Bahia, CE=Ceará, MA=Maranhão, PB=Paraíba, PE=Pernambuco, PI=Piauí, RN=Rio Grande do Norte, and SE=Sergipe; Center-West: DF=Distrito Federal, GO=Goiás, MT=Mato Grosso, and MS=Mato Grosso do Sul; Southeast: ES=Espírito Santo, MG=Minas Gerais, RJ=Rio de Janeiro, and SP=São Paulo; South: PR=Paraná, RS=Rio Grande do Sul, and SC=Santa Catarina. **(B)** Decline (in years) in life expectancy at age 65 due to COVID-19 for both sexes, **(C)** females, and **(D)** males under two alternative scenarios.

**Fig. S2.**
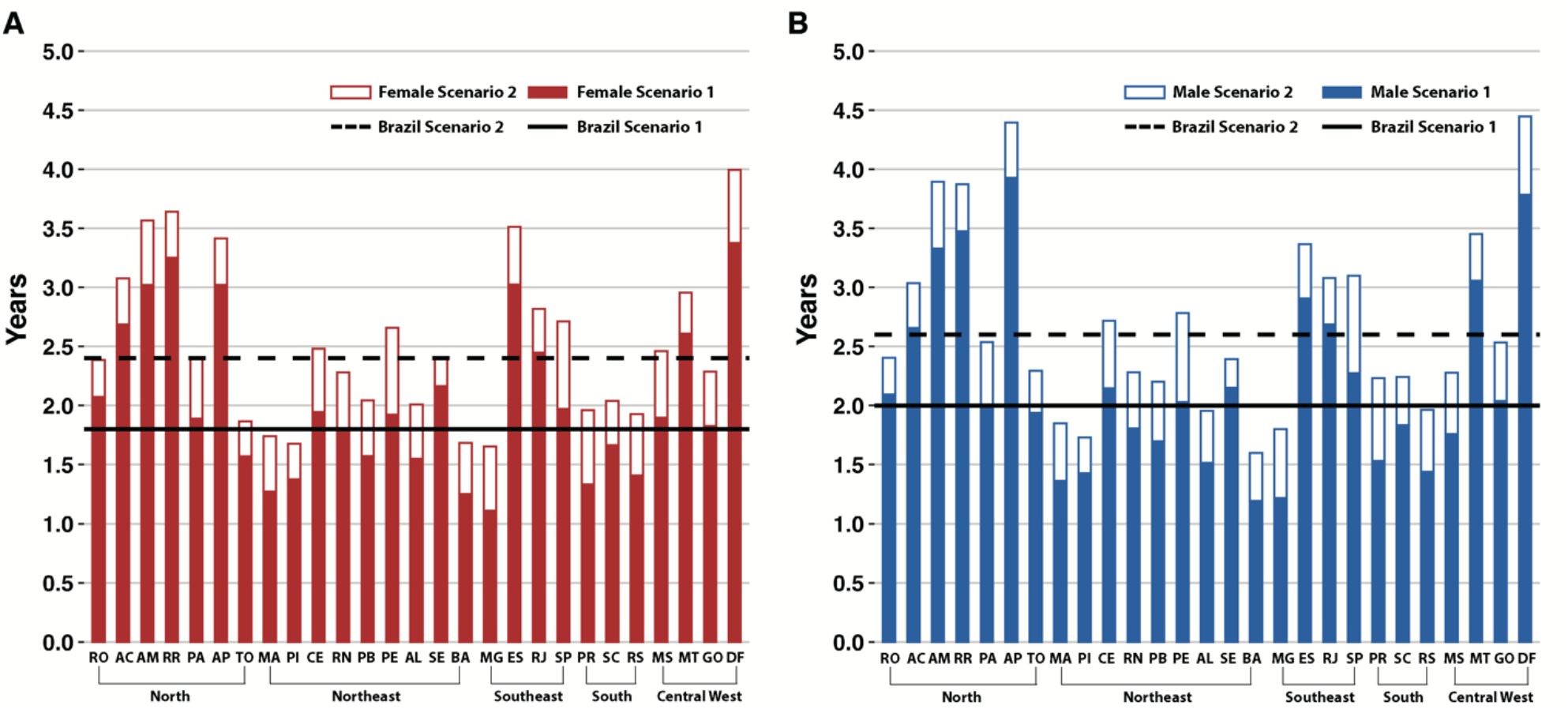
Decline in life expectancy at birth. **(A)** Decline (in years) in life expectancy at birth in 2020 for females and **(B)** males, considering COVID-19 mortality under two alternative scenarios. Scenario (1) considers all COVID-19 deaths reported in 2020. Scenario (2) assumes that, under a pandemic, 90% of the unspecified severe acute respiratory infections (SARI) deaths in each state were unidentified COVID-19 deaths (see **Materials and Methods**).

**Fig. S3.**
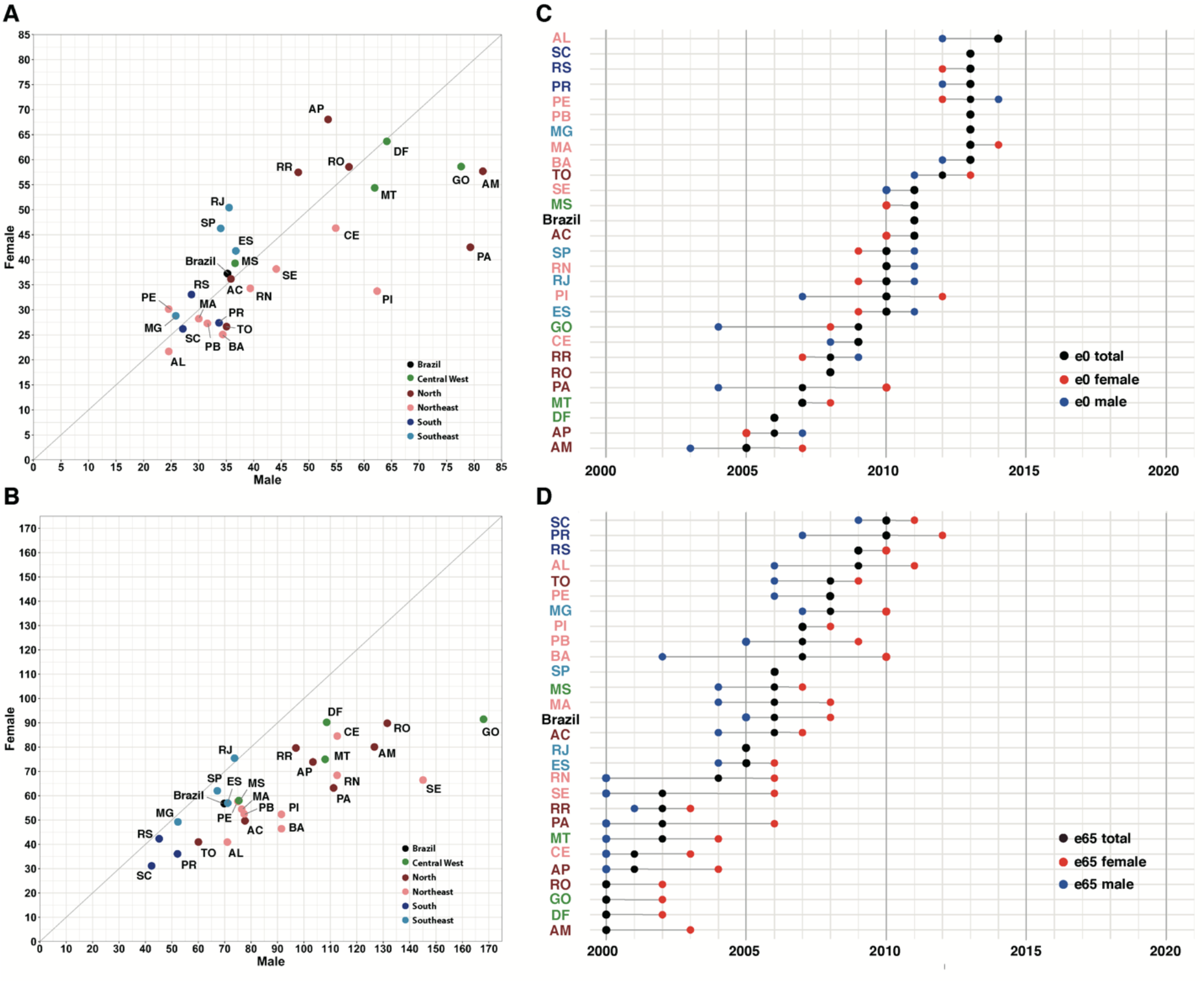
Loss and setback in life expectancy at birth and at age 65 by state and sex. Percentage loss due to COVID-19 mortality relative to increases in female and male life expectancy at birth **(A)** and age 65 **(B)** between 2000 and 2020, by state. Results for Scenario (2), which assumes that, under a pandemic, 90% of the unspecified severe acute respiratory infections (SARI) deaths in each state were unidentified COVID-19 deaths (see **Materials and Methods**). States are colored according to major regions. State acronyms by region, North: AC=Acre, AP=Amapá, AM=Amazonas, PA=Pará, RO=Rondônia, RR=Roraima, and TO=Tocantins; Northeast: AL=Alagoas, BA=Bahia, CE=Ceará, MA=Maranhão, PB=Paraíba, PE=Pernambuco, PI=Piauí, RN=Rio Grande do Norte, and SE=Sergipe; Center-West: DF=Distrito Federal, GO=Goiás, MT=Mato Grosso, and MS=Mato Grosso do Sul; Southeast: ES=Espírito Santo, MG=Minas Gerais, RJ=Rio de Janeiro, and SP=São Paulo; South: PR=Paraná, RS=Rio Grande do Sul, and SC=Santa Catarina. Setback in life expectancy at birth **(C)** and age 65 **(D)** due to COVID-19 mortality, by sex and state. State acronyms are colored according to major regions. In some states the setback is similar among total, males, or females (**Table S9**).

**Table S1.** Estimated life expectancy at birth *(e*_0_) in 2020 with and without COVID-19, and absolute and percentage decline due to COVID-19. Estimates by sex and state, based on the total number of COVID-19 deaths reported in 2020.

| Region | State | Total $e_0$ 2020 | | | | Female $e_0$ 2020 | | | | Male $e_0$ 2020 | | | |
| --- | --- | --- | --- | --- | --- | --- | --- | --- | --- | --- | --- | --- | --- |
|  |  | Without COVID | With COVID | Decline | % Decline | Without COVID | With COVID | Decline | % Decline | Without COVID | With COVID | Decline | % Decline |
| North | RO | 72.09 | 69.99 | 2.10 | 2.92 | 75.67 | 73.60 | 2.07 | 2.74 | 69.00 | 66.90 | 2.09 | 3.03 |
|  | AC | 75.09 | 72.37 | 2.72 | 3.62 | 78.60 | 75.92 | 2.69 | 3.42 | 71.87 | 69.21 | 2.66 | 3.70 |
|  | AM | 72.81 | 69.53 | 3.28 | 4.51 | 76.48 | 73.46 | 3.02 | 3.95 | 69.47 | 66.14 | 3.33 | 4.79 |
|  | RR | 72.69 | 69.26 | 3.43 | 4.72 | 75.34 | 72.08 | 3.25 | 4.32 | 70.36 | 66.88 | 3.48 | 4.94 |
|  | PA | 72.83 | 70.82 | 2.01 | 2.76 | 77.16 | 75.27 | 1.89 | 2.45 | 69.00 | 66.99 | 2.01 | 2.91 |
|  | AP | 74.88 | 71.27 | 3.62 | 4.83 | 77.52 | 74.50 | 3.02 | 3.90 | 72.41 | 68.49 | 3.93 | 5.42 |
|  | TO | 74.39 | 72.59 | 1.80 | 2.42 | 77.70 | 76.13 | 1.57 | 2.02 | 71.44 | 69.51 | 1.94 | 2.71 |
| Northeast | MA | 71.67 | 70.31 | 1.37 | 1.91 | 75.57 | 74.30 | 1.27 | 1.68 | 67.96 | 66.60 | 1.36 | 2.00 |
|  | PI | 71.76 | 70.32 | 1.45 | 2.02 | 76.17 | 74.80 | 1.38 | 1.81 | 67.47 | 66.05 | 1.43 | 2.11 |
|  | CE | 74.68 | 72.59 | 2.09 | 2.80 | 78.65 | 76.71 | 1.94 | 2.47 | 70.75 | 68.61 | 2.15 | 3.03 |
|  | RN | 76.57 | 74.73 | 1.84 | 2.41 | 80.56 | 78.76 | 1.80 | 2.23 | 72.60 | 70.79 | 1.81 | 2.49 |
|  | PB | 74.36 | 72.70 | 1.66 | 2.23 | 78.19 | 76.62 | 1.57 | 2.01 | 70.47 | 68.77 | 1.70 | 2.41 |
|  | PE | 75.31 | 73.30 | 2.01 | 2.67 | 78.92 | 76.99 | 1.92 | 2.44 | 71.55 | 69.52 | 2.03 | 2.84 |
|  | AL | 72.98 | 71.41 | 1.57 | 2.16 | 77.76 | 76.21 | 1.55 | 1.99 | 68.26 | 66.75 | 1.51 | 2.22 |
|  | SE | 73.64 | 71.43 | 2.21 | 3.00 | 77.91 | 75.75 | 2.16 | 2.78 | 69.42 | 67.27 | 2.15 | 3.10 |
|  | BA | 74.36 | 73.11 | 1.25 | 1.68 | 79.11 | 77.86 | 1.25 | 1.58 | 69.86 | 68.66 | 1.19 | 1.71 |
| Southeast | MG | 78.19 | 77.01 | 1.18 | 1.51 | 81.04 | 79.93 | 1.11 | 1.37 | 75.37 | 74.16 | 1.22 | 1.61 |
|  | ES | 79.32 | 76.30 | 3.02 | 3.80 | 83.21 | 80.18 | 3.03 | 3.64 | 75.56 | 72.66 | 2.91 | 3.85 |
|  | RJ | 77.30 | 74.68 | 2.62 | 3.39 | 80.39 | 77.94 | 2.45 | 3.04 | 73.97 | 71.29 | 2.69 | 3.63 |
|  | SP | 79.11 | 76.94 | 2.17 | 2.74 | 81.96 | 79.99 | 1.97 | 2.41 | 76.12 | 73.85 | 2.27 | 2.99 |
| South | PR | 78.21 | 76.74 | 1.46 | 1.87 | 81.65 | 80.32 | 1.33 | 1.63 | 74.83 | 73.30 | 1.53 | 2.05 |
|  | SC | 80.21 | 78.41 | 1.80 | 2.24 | 83.49 | 81.83 | 1.66 | 1.99 | 76.96 | 75.12 | 1.84 | 2.39 |
|  | RS | 78.79 | 77.34 | 1.45 | 1.84 | 82.04 | 80.63 | 1.41 | 1.71 | 75.44 | 74.00 | 1.44 | 1.91 |
| Central West | MS | 76.53 | 74.68 | 1.85 | 2.42 | 80.16 | 78.26 | 1.90 | 2.37 | 73.12 | 71.36 | 1.76 | 2.41 |
|  | MT | 75.17 | 72.27 | 2.90 | 3.86 | 78.74 | 76.13 | 2.61 | 3.31 | 72.07 | 69.02 | 3.06 | 4.24 |
|  | GO | 74.83 | 72.86 | 1.97 | 2.63 | 78.20 | 76.38 | 1.83 | 2.34 | 71.66 | 69.63 | 2.04 | 2.84 |
|  | DF | 79.08 | 75.40 | 3.68 | 4.66 | 82.38 | 79.00 | 3.37 | 4.10 | 75.43 | 71.65 | 3.79 | 5.02 |
| Brazil |  | 76.74 | 74.80 | 1.94 | 2.53 | 80.25 | 78.43 | 1.82 | 2.26 | 73.26 | 71.28 | 1.98 | 2.70 |

**Table S2.** Estimated life expectancy at age 65 *(e*_65_) in 2020 with and without COVID-19, and absolute and percentage decline due to COVID-19. Estimates by sex and state, based on the total number of COVID-19 deaths reported in 2020.

| Region | State | Total $e_{65}$ 2020 | | | | Female $e_{65}$ 2020 | | | | Male $e_{65}$ 2020 | | | |
| --- | --- | --- | --- | --- | --- | --- | --- | --- | --- | --- | --- | --- | --- |
|  |  | Without COVID | With COVID | Decline | % Decline | Without COVID | With COVID | Decline | % Decline | Without COVID | With COVID | Decline | % Decline |
| North | RO | 16.24 | 14.57 | 1.67 | 10.26 | 17.51 | 15.84 | 1.67 | 9.54 | 15.11 | 13.47 | 1.64 | 10.87 |
|  | AC | 18.51 | 16.31 | 2.20 | 11.90 | 20.19 | 18.05 | 2.13 | 10.57 | 16.85 | 14.67 | 2.17 | 12.90 |
|  | AM | 17.23 | 14.31 | 2.92 | 16.96 | 18.72 | 16.12 | 2.60 | 13.89 | 15.72 | 12.67 | 3.05 | 19.43 |
|  | RR | 16.73 | 14.00 | 2.74 | 16.35 | 17.76 | 15.19 | 2.57 | 14.45 | 15.81 | 12.99 | 2.82 | 17.82 |
|  | PA | 17.23 | 15.47 | 1.76 | 10.20 | 18.80 | 17.23 | 1.57 | 8.34 | 15.67 | 13.83 | 1.84 | 11.74 |
|  | AP | 18.51 | 15.53 | 2.98 | 16.09 | 19.75 | 17.40 | 2.36 | 11.92 | 17.25 | 13.89 | 3.36 | 19.48 |
|  | TO | 17.98 | 16.48 | 1.50 | 8.34 | 19.20 | 17.87 | 1.33 | 6.92 | 16.85 | 15.25 | 1.60 | 9.48 |
| Northeast | MA | 17.34 | 16.11 | 1.23 | 7.12 | 19.15 | 18.07 | 1.08 | 5.64 | 15.41 | 14.13 | 1.28 | 8.32 |
|  | PI | 16.54 | 15.35 | 1.20 | 7.23 | 18.10 | 16.96 | 1.14 | 6.28 | 14.75 | 13.57 | 1.18 | 8.00 |
|  | CE | 18.10 | 16.35 | 1.76 | 9.71 | 19.37 | 17.76 | 1.61 | 8.33 | 16.58 | 14.74 | 1.84 | 11.09 |
|  | RN | 18.94 | 17.48 | 1.46 | 7.70 | 20.57 | 19.16 | 1.40 | 6.83 | 16.97 | 15.52 | 1.45 | 8.55 |
|  | PB | 17.90 | 16.60 | 1.31 | 7.29 | 19.02 | 17.79 | 1.24 | 6.50 | 16.51 | 15.16 | 1.35 | 8.17 |
|  | PE | 18.02 | 16.42 | 1.60 | 8.89 | 19.32 | 17.83 | 1.49 | 7.72 | 16.33 | 14.67 | 1.66 | 10.17 |
|  | AL | 17.57 | 16.32 | 1.25 | 7.13 | 19.17 | 18.02 | 1.15 | 5.99 | 15.67 | 14.38 | 1.29 | 8.22 |
|  | SE | 17.37 | 15.64 | 1.74 | 10.00 | 18.95 | 17.33 | 1.62 | 8.55 | 15.49 | 13.73 | 1.76 | 11.38 |
|  | BA | 18.38 | 17.34 | 1.04 | 5.64 | 20.21 | 19.21 | 1.00 | 4.97 | 16.30 | 15.29 | 1.02 | 6.24 |
| Southeast | MG | 19.65 | 18.63 | 1.02 | 5.17 | 20.87 | 19.94 | 0.94 | 4.48 | 18.26 | 17.18 | 1.08 | 5.89 |
|  | ES | 20.62 | 18.17 | 2.45 | 11.89 | 22.42 | 20.04 | 2.38 | 10.62 | 18.62 | 16.19 | 2.43 | 13.04 |
|  | RJ | 19.09 | 17.05 | 2.04 | 10.70 | 20.57 | 18.68 | 1.89 | 9.20 | 17.14 | 15.02 | 2.11 | 12.33 |
|  | SP | 19.78 | 18.05 | 1.73 | 8.75 | 21.26 | 19.67 | 1.59 | 7.47 | 17.97 | 16.16 | 1.81 | 10.08 |
| South | PR | 19.28 | 18.12 | 1.16 | 6.04 | 20.79 | 19.73 | 1.06 | 5.09 | 17.63 | 16.40 | 1.23 | 6.98 |
|  | SC | 20.44 | 18.92 | 1.52 | 7.44 | 22.35 | 20.96 | 1.39 | 6.23 | 18.27 | 16.71 | 1.57 | 8.57 |
|  | RS | 19.65 | 18.45 | 1.20 | 6.09 | 21.36 | 20.21 | 1.15 | 5.38 | 17.58 | 16.39 | 1.19 | 6.78 |
| Central West | MS | 18.83 | 17.29 | 1.54 | 8.16 | 20.48 | 18.94 | 1.54 | 7.52 | 17.10 | 15.61 | 1.49 | 8.71 |
|  | MT | 18.12 | 15.92 | 2.20 | 12.15 | 19.47 | 17.47 | 2.01 | 10.30 | 16.89 | 14.58 | 2.31 | 13.69 |
|  | GO | 17.80 | 16.16 | 1.64 | 9.21 | 18.91 | 17.44 | 1.47 | 7.78 | 16.62 | 14.86 | 1.76 | 10.58 |
|  | DF | 19.72 | 16.63 | 3.08 | 15.63 | 21.38 | 18.52 | 2.86 | 13.38 | 17.66 | 14.51 | 3.15 | 17.85 |
| Brazil |  | 19.01 | 17.43 | 1.58 | 8.30 | 20.51 | 19.06 | 1.46 | 7.11 | 17.25 | 15.61 | 1.64 | 9.48 |

**Table S3.**
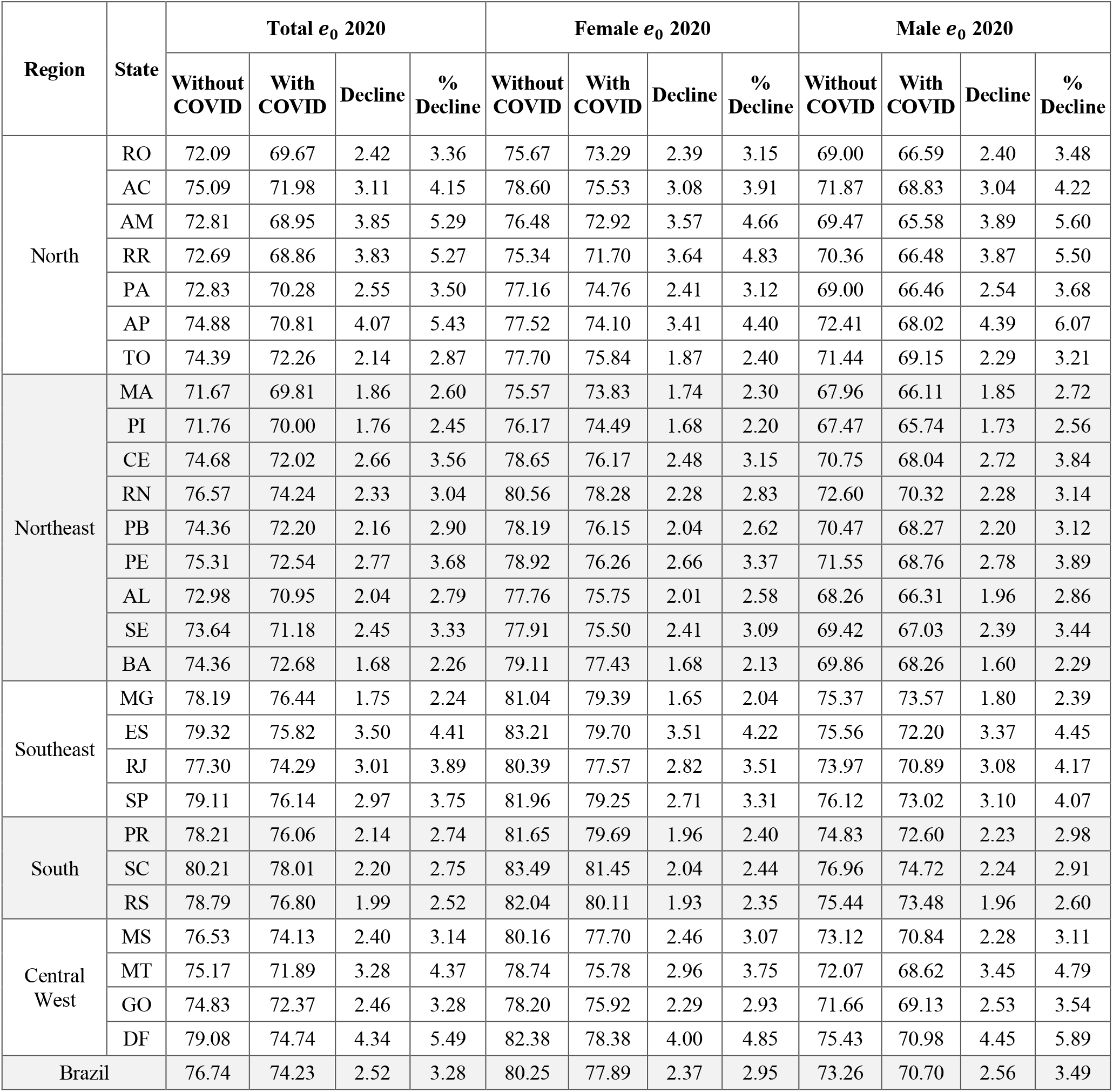
Estimated life expectancy at birth *(e*_0_) in 2020 with and without COVID-19, and absolute and percentage decline due to COVID-19. Estimates by sex and state, based on an alternative scenario of COVID-19 mortality that accounts for the misdiagnosis of severe acute respiratory infections (SARI). Specifically, we assumed that, under a pandemic, 90% of the unspecified severe acute respiratory infections (SARI) deaths in each state were unidentified COVID-19 deaths.

| Region | State | Total $e_0$ 2020 | | | | Female $e_0$ 2020 | | | | Male $e_0$ 2020 | | | |
| --- | --- | --- | --- | --- | --- | --- | --- | --- | --- | --- | --- | --- | --- |
|  |  | Without COVID | With COVID | Decline | % Decline | Without COVID | With COVID | Decline | % Decline | Without COVID | With COVID | Decline | % Decline |
| North | RO | 72.09 | 69.67 | 2.42 | 3.36 | 75.67 | 73.29 | 2.39 | 3.15 | 69.00 | 66.59 | 2.40 | 3.48 |
|  | AC | 75.09 | 71.98 | 3.11 | 4.15 | 78.60 | 75.53 | 3.08 | 3.91 | 71.87 | 68.83 | 3.04 | 4.22 |
|  | AM | 72.81 | 68.95 | 3.85 | 5.29 | 76.48 | 72.92 | 3.57 | 4.66 | 69.47 | 65.58 | 3.89 | 5.60 |
|  | RR | 72.69 | 68.86 | 3.83 | 5.27 | 75.34 | 71.70 | 3.64 | 4.83 | 70.36 | 66.48 | 3.87 | 5.50 |
|  | PA | 72.83 | 70.28 | 2.55 | 3.50 | 77.16 | 74.76 | 2.41 | 3.12 | 69.00 | 66.46 | 2.54 | 3.68 |
|  | AP | 74.88 | 70.81 | 4.07 | 5.43 | 77.52 | 74.10 | 3.41 | 4.40 | 72.41 | 68.02 | 4.39 | 6.07 |
|  | TO | 74.39 | 72.26 | 2.14 | 2.87 | 77.70 | 75.84 | 1.87 | 2.40 | 71.44 | 69.15 | 2.29 | 3.21 |
| Northeast | MA | 71.67 | 69.81 | 1.86 | 2.60 | 75.57 | 73.83 | 1.74 | 2.30 | 67.96 | 66.11 | 1.85 | 2.72 |
|  | PI | 71.76 | 70.00 | 1.76 | 2.45 | 76.17 | 74.49 | 1.68 | 2.20 | 67.47 | 65.74 | 1.73 | 2.56 |
|  | CE | 74.68 | 72.02 | 2.66 | 3.56 | 78.65 | 76.17 | 2.48 | 3.15 | 70.75 | 68.04 | 2.72 | 3.84 |
|  | RN | 76.57 | 74.24 | 2.33 | 3.04 | 80.56 | 78.28 | 2.28 | 2.83 | 72.60 | 70.32 | 2.28 | 3.14 |
|  | PB | 74.36 | 72.20 | 2.16 | 2.90 | 78.19 | 76.15 | 2.04 | 2.62 | 70.47 | 68.27 | 2.20 | 3.12 |
|  | PE | 75.31 | 72.54 | 2.77 | 3.68 | 78.92 | 76.26 | 2.66 | 3.37 | 71.55 | 68.76 | 2.78 | 3.89 |
|  | AL | 72.98 | 70.95 | 2.04 | 2.79 | 77.76 | 75.75 | 2.01 | 2.58 | 68.26 | 66.31 | 1.96 | 2.86 |
|  | SE | 73.64 | 71.18 | 2.45 | 3.33 | 77.91 | 75.50 | 2.41 | 3.09 | 69.42 | 67.03 | 2.39 | 3.44 |
|  | BA | 74.36 | 72.68 | 1.68 | 2.26 | 79.11 | 77.43 | 1.68 | 2.13 | 69.86 | 68.26 | 1.60 | 2.29 |
| Southeast | MG | 78.19 | 76.44 | 1.75 | 2.24 | 81.04 | 79.39 | 1.65 | 2.04 | 75.37 | 73.57 | 1.80 | 2.39 |
|  | ES | 79.32 | 75.82 | 3.50 | 4.41 | 83.21 | 79.70 | 3.51 | 4.22 | 75.56 | 72.20 | 3.37 | 4.45 |
|  | RJ | 77.30 | 74.29 | 3.01 | 3.89 | 80.39 | 77.57 | 2.82 | 3.51 | 73.97 | 70.89 | 3.08 | 4.17 |
|  | SP | 79.11 | 76.14 | 2.97 | 3.75 | 81.96 | 79.25 | 2.71 | 3.31 | 76.12 | 73.02 | 3.10 | 4.07 |
| South | PR | 78.21 | 76.06 | 2.14 | 2.74 | 81.65 | 79.69 | 1.96 | 2.40 | 74.83 | 72.60 | 2.23 | 2.98 |
|  | SC | 80.21 | 78.01 | 2.20 | 2.75 | 83.49 | 81.45 | 2.04 | 2.44 | 76.96 | 74.72 | 2.24 | 2.91 |
|  | RS | 78.79 | 76.80 | 1.99 | 2.52 | 82.04 | 80.11 | 1.93 | 2.35 | 75.44 | 73.48 | 1.96 | 2.60 |
| Central West | MS | 76.53 | 74.13 | 2.40 | 3.14 | 80.16 | 77.70 | 2.46 | 3.07 | 73.12 | 70.84 | 2.28 | 3.11 |
|  | MT | 75.17 | 71.89 | 3.28 | 4.37 | 78.74 | 75.78 | 2.96 | 3.75 | 72.07 | 68.62 | 3.45 | 4.79 |
|  | GO | 74.83 | 72.37 | 2.46 | 3.28 | 78.20 | 75.92 | 2.29 | 2.93 | 71.66 | 69.13 | 2.53 | 3.54 |
|  | DF | 79.08 | 74.74 | 4.34 | 5.49 | 82.38 | 78.38 | 4.00 | 4.85 | 75.43 | 70.98 | 4.45 | 5.89 |
| Brazil |  | 76.74 | 74.23 | 2.52 | 3.28 | 80.25 | 77.89 | 2.37 | 2.95 | 73.26 | 70.70 | 2.56 | 3.49 |

**Table S4.**
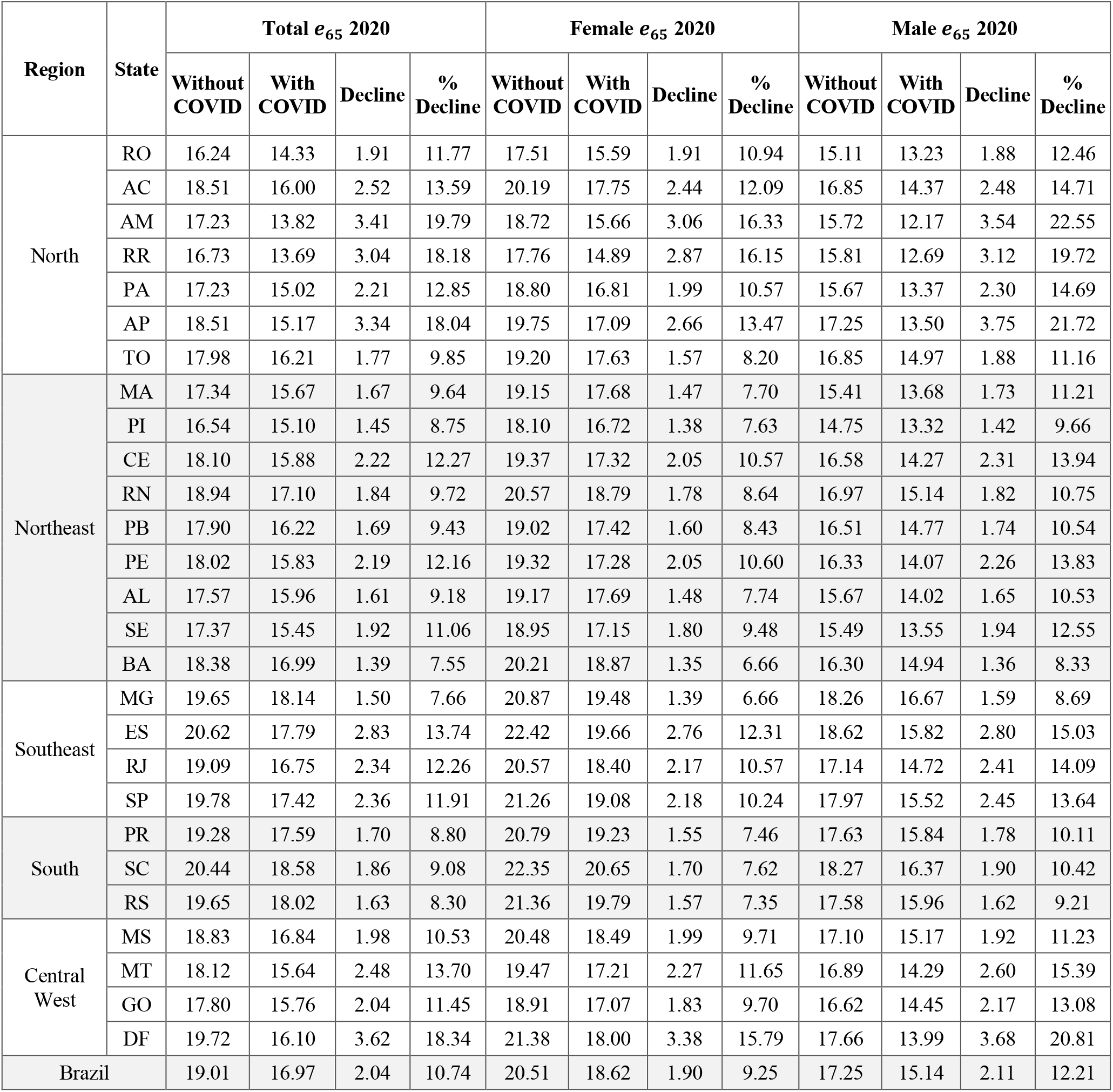
Estimated life expectancy at age 65 *(e*_65_) in 2020 with and without COVID-19, and absolute and percentage decline due to COVID-19. Estimates by sex and state, based on an alternative scenario of COVID-19 mortality that accounts for the misdiagnosis of severe acute respiratory infections (SARI). Specifically, we assumed that, under a pandemic, 90% of the unspecified severe acute respiratory infections (SARI) deaths in each state were unidentified COVID-19 deaths.

**Table S5.** Estimated gap in female-male life expectancy at birth *(e*_0_) and at age 65 *(e*_65_) in 2020 with and without COVID-19, under two alternative scenarios, by state. Scenario (1) is based on the total number of COVID-19 deaths reported in 2020. Scenario (2) accounts for the misdiagnosis of severe acute respiratory infections (SARI). Specifically, we assumed that, under a pandemic, 90% of the unspecified severe acute respiratory infections (SARI) deaths in each state were unidentified COVID-19 deaths.

| Region | State | $e_0$ | | | | | $e_{65}$ | | | | |
| --- | --- | --- | --- | --- | --- | --- | --- | --- | --- | --- | --- |
|  |  | Gap 2020 | Gap (1) | Change (1) | Gap (2) | Change (2) | Gap 2020 | Gap (1) | Change (1) | Gap (2) | Change (2) |
| North | RO | 6.68 | 6.70 | 0.02 | 6.69 | 0.02 | 2.39 | 2.36 | -0.03 | 2.36 | -0.03 |
|  | AC | 6.74 | 6.71 | -0.03 | 6.70 | -0.04 | 3.34 | 3.38 | 0.04 | 3.38 | 0.04 |
|  | AM | 7.01 | 7.32 | 0.31 | 7.34 | 0.33 | 3.00 | 3.45 | 0.45 | 3.49 | 0.49 |
|  | RR | 4.98 | 5.20 | 0.22 | 5.21 | 0.23 | 1.95 | 2.20 | 0.25 | 2.20 | 0.25 |
|  | PA | 8.16 | 8.28 | 0.12 | 8.29 | 0.13 | 3.13 | 3.40 | 0.27 | 3.44 | 0.32 |
|  | AP | 5.10 | 6.01 | 0.91 | 6.08 | 0.98 | 2.50 | 3.51 | 1.01 | 3.59 | 1.09 |
|  | TO | 6.26 | 6.63 | 0.37 | 6.69 | 0.43 | 2.35 | 2.62 | 0.27 | 2.66 | 0.31 |
| Northeast | MA | 7.61 | 7.70 | 0.09 | 7.72 | 0.11 | 3.74 | 3.94 | 0.20 | 3.99 | 0.25 |
|  | PI | 8.70 | 8.75 | 0.05 | 8.75 | 0.05 | 3.35 | 3.39 | 0.04 | 3.39 | 0.04 |
|  | CE | 7.90 | 8.10 | 0.20 | 8.14 | 0.24 | 2.79 | 3.02 | 0.23 | 3.05 | 0.26 |
|  | RN | 7.96 | 7.97 | 0.01 | 7.96 | 0.00 | 3.60 | 3.65 | 0.05 | 3.65 | 0.05 |
|  | PB | 7.72 | 7.85 | 0.13 | 7.88 | 0.16 | 2.51 | 2.63 | 0.11 | 2.65 | 0.14 |
|  | PE | 7.37 | 7.48 | 0.11 | 7.49 | 0.13 | 2.99 | 3.16 | 0.17 | 3.20 | 0.21 |
|  | AL | 9.50 | 9.47 | -0.04 | 9.45 | -0.05 | 3.50 | 3.64 | 0.14 | 3.67 | 0.17 |
|  | SE | 8.48 | 8.47 | 0.01 | 8.47 | -0.02 | 3.46 | 3.60 | 0.14 | 3.61 | 0.15 |
|  | BA | 9.26 | 9.20 | -0.06 | 9.17 | -0.08 | 3.91 | 3.92 | 0.01 | 3.92 | 0.01 |
| Southeast | MG | 5.67 | 5.78 | 0.11 | 5.82 | 0.15 | 2.61 | 2.75 | 0.14 | 2.81 | 0.20 |
|  | ES | 7.65 | 7.53 | -0.12 | 7.50 | -0.15 | 3.80 | 3.85 | 0.05 | 3.84 | 0.04 |
|  | RJ | 6.41 | 6.65 | 0.24 | 6.68 | 0.26 | 3.43 | 3.65 | 0.22 | 3.67 | 0.24 |
|  | SP | 5.84 | 6.14 | 0.30 | 6.23 | 0.39 | 3.29 | 3.51 | 0.22 | 3.56 | 0.28 |
| South | PR | 6.83 | 7.02 | 0.20 | 7.10 | 0.27 | 3.16 | 3.33 | 0.17 | 3.39 | 0.23 |
|  | SC | 6.53 | 6.70 | 0.17 | 6.73 | 0.20 | 4.08 | 4.25 | 0.17 | 4.28 | 0.20 |
|  | RS | 6.59 | 6.63 | 0.03 | 6.63 | 0.03 | 3.78 | 3.82 | 0.04 | 3.83 | 0.05 |
| Central West | MS | 7.04 | 6.90 | -0.14 | 6.86 | -0.18 | 3.39 | 3.34 | -0.05 | 3.32 | -0.07 |
|  | MT | 6.67 | 7.11 | 0.45 | 7.16 | 0.50 | 2.58 | 2.89 | 0.31 | 2.91 | 0.33 |
|  | GO | 6.54 | 6.75 | 0.21 | 6.79 | 0.25 | 2.28 | 2.57 | 0.29 | 2.62 | 0.34 |
|  | DF | 6.95 | 7.36 | 0.41 | 7.40 | 0.45 | 3.71 | 4.00 | 0.29 | 4.01 | 0.30 |
| Brazil |  | 6.99 | 7.15 | 2.32 | 0.16 | 7.18 | 0.19 | 3.27 | 3.45 | 0.18 | 3.48 |

**Table S6.** Regional gap (considering the extremes in state life expectancy) in life expectancy at birth *(e*_0_) and at age 65 *(e*_65_) from 2000 to 2019, and estimated gap in 2020 with and without COVID-19, under two alternative scenarios, by sex. Scenario (1) is based on the total number of COVID-19 deaths reported in 2020. Scenario (2) accounts for the misdiagnosis of severe acute respiratory infections (SARI). Specifically, we assumed that, under a pandemic, 90% of the unspecified severe acute respiratory infections (SARI) deaths in each state were unidentified COVID-19 deaths.

| Year | $e_0$ | | | | | | $e_{65}$ | | | | | |
| --- | --- | --- | --- | --- | --- | --- | --- | --- | --- | --- | --- | --- |
|  | Total | States<br>(High, Low) | Female | States<br>(High, Low) | Male | States<br>(High, Low) | Total | States<br>(High, Low) | Female | States<br>(High, Low) | Male | States<br>(High, Low) |
| 2000 | 8.10 | (RS, AL) | 7.70 | (RS, AL) | 8.50 | (SC, PE) | 3.42 | (MG, RR) | 3.89 | (MG, RR) | 2.75 | (RN, RR) |
| 2001 | 8.00 | (RS, AL) | 7.50 | (RS, AL) | 8.50 | (SC, AL) | 3.40 | (MG, RR) | 3.84 | (MG, RR) | 2.67 | (MG, RR) |
| 2002 | 7.90 | (RS/SC, AL) | 7.20 | (RS/DF, AL) | 8.60 | (SC, AL) | 3.39 | (MG, RR) | 3.78 | (MG, RR) | 2.70 | (MG, RR) |
| 2003 | 7.90 | (SC, AL) | 7.30 | (DF, RR) | 8.60 | (SC, AL) | 3.37 | (MG, RR) | 3.81 | (ES, RR) | 2.78 | (MG, PI) |
| 2004 | 7.90 | (SC, AL) | 7.30 | (DF, RR) | 8.70 | (SC, AL) | 3.35 | (MG, RR) | 3.93 | (ES, RR) | 3.05 | (MG, PI) |
| 2005 | 7.90 | (SC, AL) | 7.40 | (DF/SC, RR) | 8.80 | (SC, AL) | 3.44 | (ES, RR) | 4.05 | (ES, RR) | 3.26 | (MG, PI) |
| 2006 | 7.90 | (SC, AL) | 7.50 | (SC, RR) | 8.80 | (SC, AL) | 3.54 | (ES, RR) | 4.15 | (ES, RR) | 3.29 | (MG, PI) |
| 2007 | 7.80 | (SC, AL) | 7.60 | (SC, RR) | 8.90 | (SC, AL) | 3.64 | (ES, RR) | 4.26 | (ES, RR) | 3.25 | (MG, PI) |
| 2008 | 8.00 | (SC, MA) | 7.60 | (SC, RR) | 8.90 | (SC, AL) | 3.72 | (ES, RR) | 4.35 | (ES, RR) | 3.17 | (ES, PI) |
| 2009 | 8.10 | (SC, MA) | 7.80 | (SC, RR) | 9.00 | (SC, AL) | 3.81 | (ES, RR) | 4.44 | (ES, RR) | 3.08 | (ES, PI) |
| 2010 | 8.20 | (SC, MA) | 7.90 | (SC, RR) | 9.00 | (SC, AL) | 3.89 | (ES, RR) | 4.53 | (ES, RR) | 3.01 | (ES, RO) |
| 2011 | 8.20 | (SC, MA) | 7.90 | (SC, RR) | 9.00 | (SC, AL) | 3.91 | (ES, RR) | 4.57 | (ES, RR) | 3.12 | (ES, PI) |
| 2012 | 8.30 | (SC, MA) | 8.00 | (SC, RR) | 9.00 | (SC, AL) | 4.00 | (ES, RO) | 4.61 | (ES, RR) | 3.23 | (ES, PI) |
| 2013 | 8.40 | (SC, MA) | 8.00 | (SC, RR) | 8.90 | (SC, AL) | 4.07 | (ES, RO) | 4.64 | (ES, RR) | 3.34 | (ES, PI) |
| 2014 | 8.40 | (SC, MA) | 8.10 | (SC, RR) | 8.90 | (SC, AL) | 4.14 | (ES, RO) | 4.66 | (ES, RR) | 3.43 | (ES, PI) |
| 2015 | 8.40 | (SC, MA) | 8.10 | (SC, RR) | 8.90 | (SC, AL) | 4.20 | (ES, RO) | 4.68 | (ES, RO) | 3.52 | (ES, PI) |
| 2016 | 8.50 | (SC, MA) | 8.10 | (SC, RR) | 8.90 | (SC, AL/MA/PI) | 4.25 | (ES, RO) | 4.74 | (ES, RO) | 3.60 | (ES, PI) |
| 2017 | 8.52 | (SC, MA) | 8.13 | (SC, RR) | 9.02 | (SC, PI) | 4.29 | (ES, RO) | 4.80 | (ES, RO) | 3.68 | (ES, PI) |
| 2018 | 8.53 | (SC, MA) | 8.14 | (SC, RR) | 9.19 | (SC, PI) | 4.33 | (ES, RO) | 4.84 | (ES, RO) | 3.75 | (ES, PI) |
| 2019 | 8.53 | (SC, MA) | 8.15 | (SC, RR) | 9.34 | (SC, PI) | 4.36 | (ES, RO) | 4.88 | (ES, RO) | 3.81 | (ES, PI) |
| 2020 | 8.54 | (SC, MA) | 8.15 | (SC, RR) | 9.48 | (SC, PI) | 4.38 | (ES, RO) | 4.91 | (ES, RO) | 3.87 | (ES, PI) |
| 2020 (1) | 9.15 | (SC, RR) | 9.74 | (SC, RR) | 9.08 | (SC, PI) | 4.92 | (SC, RR) | 5.77 | (SC, RR) | 4.52 | (MG, AM) |
| 2020 (2) | 9.14 | (SC, RR) | 9.75 | (SC, RR) | 9.14 | (SC, AM) | 4.89 | (SC, RR) | 5.76 | (SC, RR) | 4.50 | (MG, AM) |

**Table S7.**
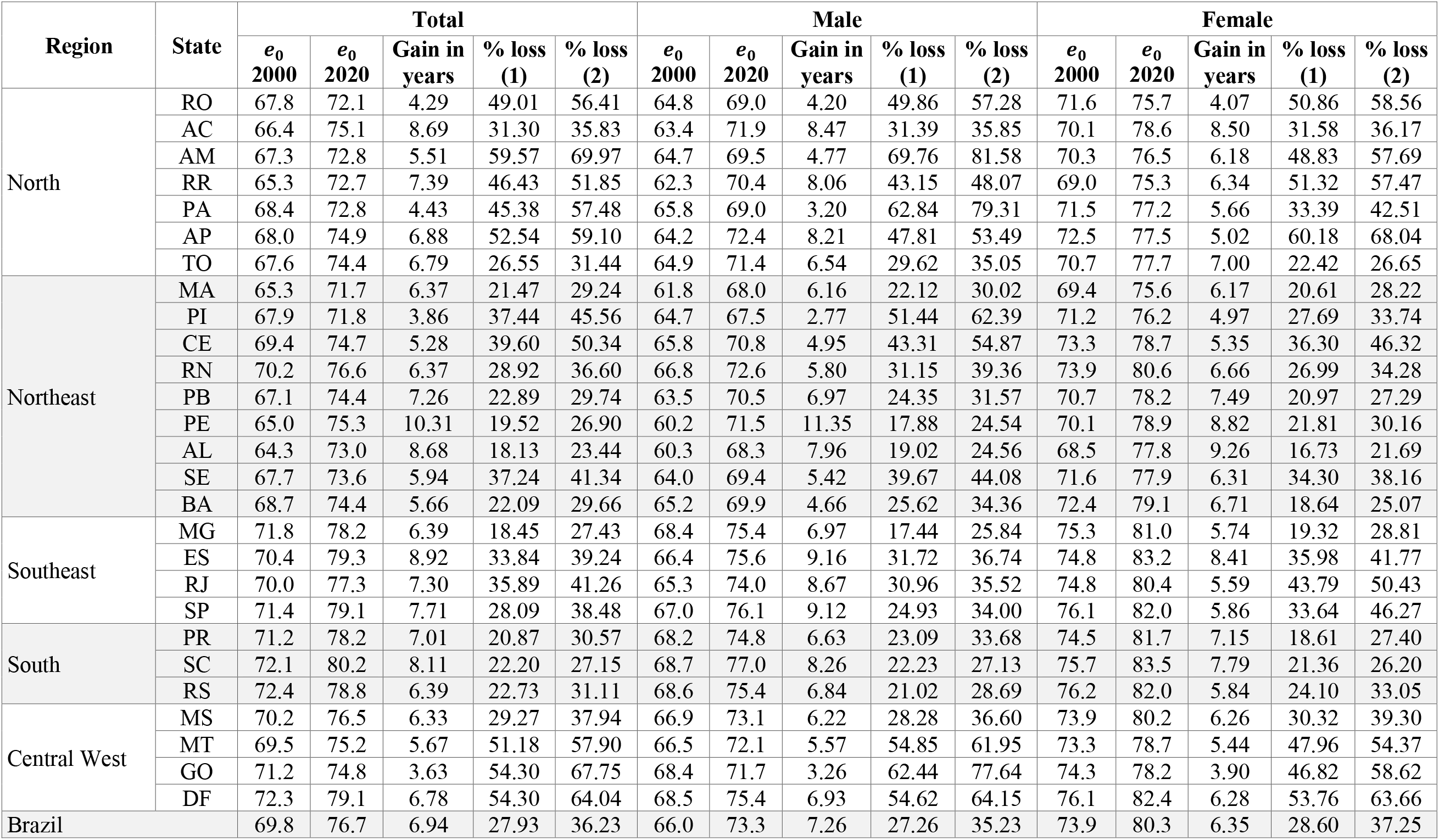
Increase in life expectancy at birth *(e*_0_) from 2000 to 2020, and estimated percentage loss of that increase due to COVID-19 under two alternative scenarios. Estimates by sex and state. Scenario (1) is based on the total number of COVID-19 deaths reported in 2020. Scenario (2) accounts for the misdiagnosis of severe acute respiratory infections (SARI). Specifically, we assumed that, under a pandemic, 90% of the unspecified severe acute respiratory infections (SARI) deaths in each state were unidentified COVID-19 deaths.

**Table S8.** Increase in life expectancy at age 65 *(e*_65_) from 2000 to 2020, and estimated percentage loss of that increase due to COVID-19 under two alternative scenarios. Estimates by sex and state. Scenario (1) is based on the total number of COVID-19 deaths reported in 2020. Scenario (2) accounts for the misdiagnosis of severe acute respiratory infections (SARI). Specifically, we assumed that, under a pandemic, 90% of the unspecified severe acute respiratory infections (SARI) deaths in each state were unidentified COVID-19 deaths.

| Region | State | Total |  |  |  |  | Male |  |  |  |  | Female |  |  |  |  |
| --- | --- | --- | --- | --- | --- | --- | --- | --- | --- | --- | --- | --- | --- | --- | --- | --- |
| | | $e_{65}$<br>2000 | $e_{65}$<br>2020 | Gain in<br>years | % loss<br>(1) | % loss<br>(2) | $e_{65}$<br>2000 | $e_{65}$<br>2020 | Gain in<br>years | % loss<br>(1) | % loss<br>(2) | $e_{65}$<br>2000 | $e_{65}$<br>2020 | Gain in<br>years | % loss<br>(1) | % loss<br>(2) |
| North | RO | 14.4 | 16.2 | 1.88 | 88.69 | 101.72 | 13.7 | 15.1 | 1.43 | 114.79 | 131.54 | 15.4 | 17.5 | 2.13 | 78.37 | 89.82 |
|  | AC | 14.4 | 18.5 | 4.15 | 53.09 | 60.64 | 13.7 | 16.8 | 3.19 | 68.12 | 77.68 | 15.3 | 20.2 | 4.91 | 43.41 | 49.68 |
|  | AM | 13.9 | 17.2 | 3.37 | 86.58 | 101.06 | 12.9 | 15.7 | 2.80 | 109.13 | 126.67 | 14.9 | 18.7 | 3.82 | 68.11 | 80.06 |
|  | RR | 13.3 | 16.7 | 3.45 | 79.39 | 88.29 | 12.6 | 15.8 | 3.22 | 87.60 | 96.96 | 14.2 | 17.8 | 3.60 | 71.23 | 79.63 |
|  | PA | 14.6 | 17.2 | 2.63 | 66.94 | 84.26 | 13.6 | 15.7 | 2.07 | 88.86 | 111.22 | 15.7 | 18.8 | 3.14 | 49.87 | 63.19 |
|  | AP | 14.8 | 18.5 | 3.66 | 81.30 | 91.14 | 13.6 | 17.3 | 3.62 | 92.75 | 103.41 | 16.2 | 19.8 | 3.60 | 65.42 | 73.88 |
|  | TO | 14.4 | 18.0 | 3.53 | 42.45 | 50.13 | 13.7 | 16.9 | 3.13 | 51.05 | 60.06 | 15.4 | 19.2 | 3.85 | 34.53 | 40.96 |
| Northeast | MA | 14.6 | 17.3 | 2.69 | 45.82 | 62.07 | 13.2 | 15.4 | 2.26 | 56.76 | 76.45 | 16.4 | 19.1 | 2.71 | 39.93 | 54.46 |
|  | PI | 14.3 | 16.5 | 2.23 | 53.64 | 64.97 | 13.2 | 14.7 | 1.56 | 75.76 | 91.47 | 15.5 | 18.1 | 2.64 | 43.11 | 52.33 |
|  | CE | 15.8 | 18.1 | 2.34 | 75.14 | 94.94 | 14.5 | 16.6 | 2.05 | 89.61 | 112.62 | 16.9 | 19.4 | 2.42 | 66.57 | 84.54 |
|  | RN | 16.7 | 18.9 | 2.28 | 63.87 | 80.55 | 15.3 | 17.0 | 1.62 | 89.51 | 112.56 | 18.0 | 20.6 | 2.60 | 54.08 | 68.42 |
|  | PB | 15.2 | 17.9 | 2.74 | 47.66 | 61.60 | 14.3 | 16.5 | 2.25 | 60.02 | 77.36 | 16.0 | 19.0 | 3.05 | 40.51 | 52.51 |
|  | PE | 14.6 | 18.0 | 3.41 | 46.99 | 64.25 | 13.3 | 16.3 | 3.01 | 55.21 | 75.03 | 15.8 | 19.3 | 3.55 | 41.97 | 57.68 |
|  | AL | 14.5 | 17.6 | 3.10 | 40.38 | 51.98 | 13.3 | 15.7 | 2.32 | 55.42 | 71.01 | 15.5 | 19.2 | 3.63 | 31.67 | 40.92 |
|  | SE | 15.3 | 17.4 | 2.11 | 82.19 | 90.97 | 14.2 | 15.5 | 1.34 | 131.48 | 145.09 | 16.2 | 19.0 | 2.70 | 59.95 | 66.47 |
|  | BA | 16.1 | 18.4 | 2.28 | 45.49 | 60.84 | 14.8 | 16.3 | 1.48 | 68.57 | 91.49 | 17.3 | 20.2 | 2.90 | 34.62 | 46.42 |
| Southeast | MG | 16.7 | 19.6 | 2.94 | 34.56 | 51.15 | 15.2 | 18.3 | 3.03 | 35.48 | 52.29 | 18.0 | 20.9 | 2.82 | 33.13 | 49.23 |
|  | ES | 16.2 | 20.6 | 4.46 | 55.01 | 63.56 | 14.7 | 18.6 | 3.93 | 61.77 | 71.17 | 17.6 | 22.4 | 4.84 | 49.19 | 56.98 |
|  | RJ | 16.0 | 19.1 | 3.11 | 65.60 | 75.16 | 13.9 | 17.1 | 3.27 | 64.54 | 73.72 | 17.7 | 20.6 | 2.88 | 65.69 | 75.45 |
|  | SP | 16.1 | 19.8 | 3.63 | 47.58 | 64.82 | 14.3 | 18.0 | 3.65 | 49.63 | 67.19 | 17.7 | 21.3 | 3.51 | 45.30 | 62.04 |
| South | PR | 15.4 | 19.3 | 3.92 | 29.69 | 43.29 | 14.2 | 17.6 | 3.42 | 35.98 | 52.14 | 16.5 | 20.8 | 4.30 | 24.61 | 36.08 |
|  | SC | 15.4 | 20.4 | 5.04 | 30.18 | 36.84 | 13.8 | 18.3 | 4.50 | 34.78 | 42.32 | 16.9 | 22.4 | 5.47 | 25.46 | 31.16 |
|  | RS | 16.0 | 19.6 | 3.65 | 32.76 | 44.67 | 14.0 | 17.6 | 3.58 | 33.26 | 45.20 | 17.6 | 21.4 | 3.71 | 30.96 | 42.31 |
| Central West | MS | 15.7 | 18.8 | 3.10 | 49.50 | 63.88 | 14.5 | 17.1 | 2.55 | 58.44 | 75.35 | 17.1 | 20.5 | 3.43 | 44.87 | 57.99 |
|  | MT | 15.3 | 18.1 | 2.78 | 79.34 | 89.45 | 14.5 | 16.9 | 2.41 | 96.07 | 108.02 | 16.4 | 19.5 | 3.02 | 66.33 | 74.98 |
|  | GO | 16.1 | 17.8 | 1.69 | 96.96 | 120.45 | 15.3 | 16.6 | 1.29 | 135.87 | 167.99 | 16.9 | 18.9 | 2.01 | 73.29 | 91.47 |
|  | DF | 16.0 | 19.7 | 3.67 | 83.93 | 98.45 | 14.3 | 17.7 | 3.38 | 93.17 | 108.62 | 17.6 | 21.4 | 3.74 | 76.44 | 90.20 |
| Brazil |  | 15.8 | 19.0 | 3.25 | 48.56 | 62.85 | 14.2 | 17.2 | 3.02 | 54.23 | 69.82 | 17.2 | 20.5 | 3.34 | 43.69 | 56.80 |

**Table S9.** Setback in 2020 life expectancy at birth *(e*_0_) and at age 65 *(e*_65_) due to COVID-19 under two alternative scenarios, by sex and state. Scenario (1) is based on the total number of COVID-19 deaths reported in 2020. Scenario (2) accounts for the misdiagnosis of severe acute respiratory infections (SARI). Specifically, we assumed that, under a pandemic, 90% of the unspecified severe acute respiratory infections (SARI) deaths in each state were unidentified COVID-19 deaths. Colors in the table represent the intensity of the setback, from low (dark yellows) to high (reds).

| Region | State | $e_0$ (1) Total | $e_0$ (1) Female | $e_0$ (1) Male | $e_0$ (2) Total | $e_0$ (2) Female | $e_0$ (2) Male | $e_{65}$ (1) Total | $e_{65}$ (1) Female | $e_{65}$ (1) Male | $e_{65}$ (2) Total | $e_{65}$ (2) Female | $e_{65}$ (2) Male |
| --- | --- | --- | --- | --- | --- | --- | --- | --- | --- | --- | --- | --- | --- |
| North | RO | 2009 | 2009 | 2009 | 2008 | 2008 | 2008 | 2002 | 2004 | 2000* | 2000* | 2002 | 2000* |
|  | AC | 2012 | 2011 | 2012 | 2011 | 2010 | 2011 | 2007 | 2008 | 2005 | 2006 | 2007 | 2004 |
|  | AM | 2007 | 2009 | 2006 | 2005 | 2007 | 2003 | 2002 | 2005 | 2000* | 2000* | 2003 | 2000* |
|  | RR | 2009 | 2009 | 2010 | 2008 | 2007 | 2009 | 2004 | 2005 | 2002 | 2002 | 2003 | 2001 |
|  | PA | 2010 | 2012 | 2007 | 2007 | 2010 | 2004 | 2005 | 2008 | 2002 | 2002 | 2006 | 2000* |
|  | AP | 2008 | 2007 | 2008 | 2006 | 2005 | 2007 | 2003 | 2005 | 2001 | 2001 | 2004 | 2000* |
|  | TO | 2013 | 2014 | 2013 | 2012 | 2013 | 2011 | 2009 | 2011 | 2007 | 2008 | 2009 | 2006 |
| Northeast | MA | 2015 | 2015 | 2015 | 2013 | 2014 | 2013 | 2009 | 2010 | 2007 | 2006 | 2008 | 2004 |
|  | PI | 2012 | 2014 | 2010 | 2010 | 2012 | 2007 | 2008 | 2010 | 2008 | 2007 | 2008 | 2007 |
|  | CE | 2011 | 2011 | 2010 | 2009 | 2009 | 2008 | 2004 | 2006 | 2002 | 2001 | 2003 | 2000* |
|  | RN | 2012 | 2012 | 2012 | 2010 | 2010 | 2011 | 2007 | 2009 | 2002 | 2004 | 2006 | 2000* |
|  | PB | 2014 | 2014 | 2014 | 2013 | 2013 | 2013 | 2010 | 2012 | 2008 | 2007 | 2009 | 2005 |
|  | PE | 2015 | 2014 | 2015 | 2013 | 2012 | 2014 | 2011 | 2012 | 2010 | 2008 | 2008 | 2006 |
|  | AL | 2015 | 2015 | 2016 | 2014 | 2014 | 2012 | 2011 | 2013 | 2008 | 2009 | 2011 | 2006 |
|  | SE | 2011 | 2012 | 2011 | 2011 | 2011 | 2010 | 2004 | 2008 | 2000* | 2002 | 2006 | 2000* |
|  | BA | 2015 | 2015 | 2014 | 2013 | 2013 | 2012 | 2009 | 2012 | 2006 | 2007 | 2010 | 2002 |
| Southeast | MG | 2015 | 2015 | 2015 | 2013 | 2013 | 2013 | 2011 | 2013 | 2010 | 2008 | 2010 | 2007 |
|  | ES | 2011 | 2010 | 2012 | 2010 | 2009 | 2011 | 2006 | 2008 | 2005 | 2005 | 2006 | 2004 |
|  | RJ | 2011 | 2010 | 2012 | 2010 | 2009 | 2011 | 2006 | 2007 | 2007 | 2005 | 2005 | 2005 |
|  | SP | 2012 | 2012 | 2013 | 2010 | 2009 | 2011 | 2009 | 2010 | 2009 | 2006 | 2006 | 2006 |
| South | PR | 2015 | 2015 | 2015 | 2013 | 2013 | 2012 | 2013 | 2014 | 2010 | 2010 | 2012 | 2007 |
|  | SC | 2014 | 2014 | 2014 | 2013 | 2013 | 2013 | 2012 | 2013 | 2010 | 2010 | 2011 | 2009 |
|  | RS | 2014 | 2014 | 2015 | 2013 | 2012 | 2013 | 2012 | 2013 | 2011 | 2009 | 2010 | 2009 |
| Central West | MS | 2013 | 2012 | 2013 | 2011 | 2010 | 2011 | 2009 | 2009 | 2007 | 2006 | 2007 | 2004 |
|  | MT | 2009 | 2009 | 2008 | 2007 | 2008 | 2007 | 2003 | 2006 | 2001 | 2002 | 2004 | 2000* |
|  | GO | 2009 | 2010 | 2007 | 2009 | 2008 | 2004 | 2001 | 2005 | 2000* | 2000* | 2002 | 2000* |
|  | DF | 2007 | 2007 | 2008 | 2006 | 2006 | 2006 | 2002 | 2004 | 2001 | 2000* | 2002 | 2000* |
| Brazil |  | 2013 | 2013 | 2013 | 2011 | 2011 | 2011 | 2009 | 2010 | 2008 | 2006 | 2008 | 2005 |

\* Year is pre-2000 levels.

**Table S10.**
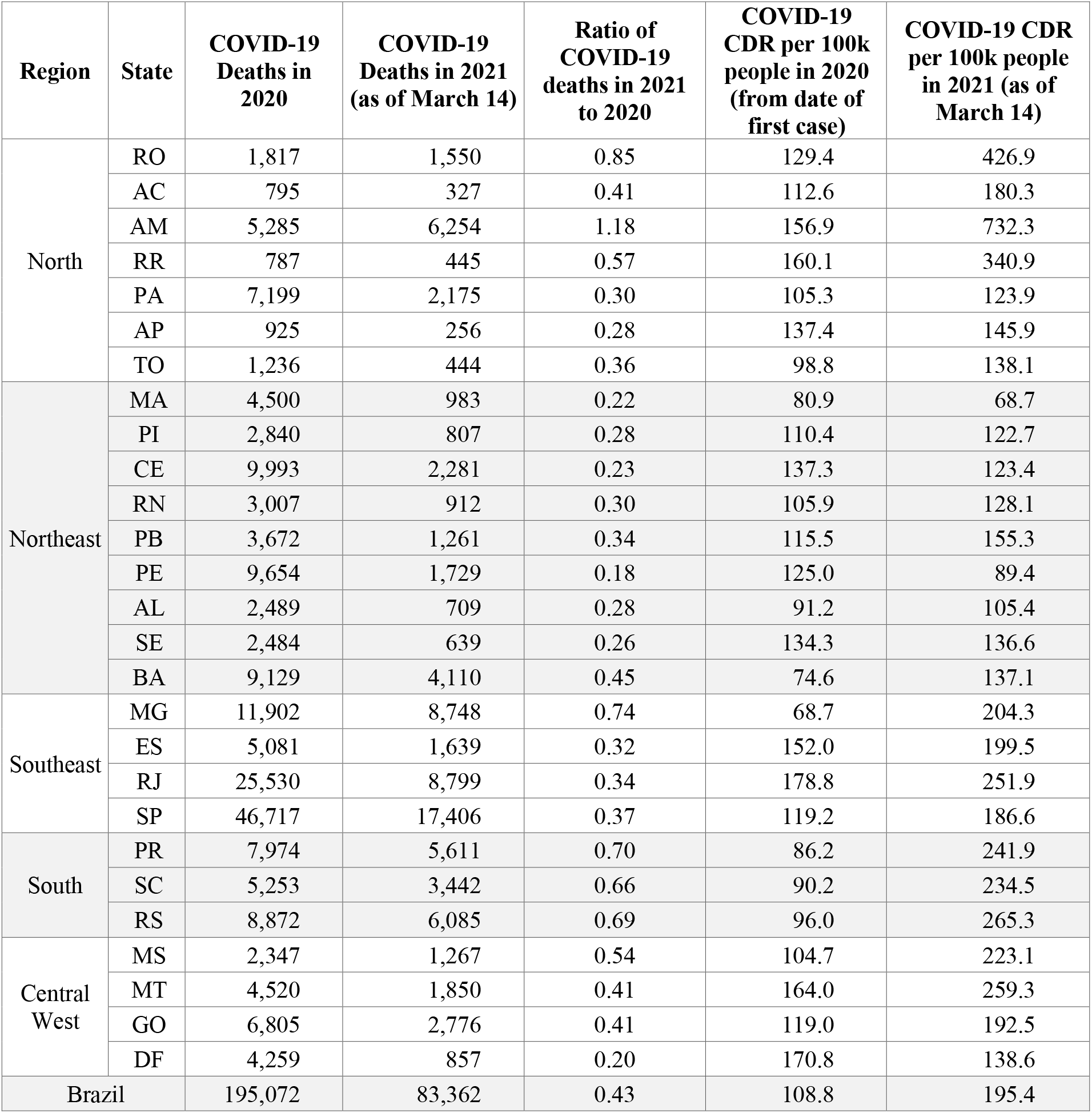
Reported COVID-19 deaths in 2020 and 2021 (as of March 14), ratio of deaths in 2021 to those in 2020, and estimated crude death rate (CDR) per 100,000 people associated with COVID-19 in 2020 and 2021, adjusting for time of exposure to risk of a COVID-19 death. In 2020, we measured exposure time as the number of days remaining in the year after the first reported case in the state. In 2021, the exposure time was measured from January 1 through March 14.

| Region | State | COVID-19 Deaths in 2020 | COVID-19 Deaths in 2021 (as of March 14) | Ratio of COVID-19 deaths in 2021 to 2020 | COVID-19 CDR per 100k people in 2020 (from date of first case) | COVID-19 CDR per 100k people in 2021 (as of March 14) |
| --- | --- | --- | --- | --- | --- | --- |
| North | RO | 1,817 | 1,550 | 0.85 | 129.4 | 426.9 |
|  | AC | 795 | 327 | 0.41 | 112.6 | 180.3 |
|  | AM | 5,285 | 6,254 | 1.18 | 156.9 | 732.3 |
|  | RR | 787 | 445 | 0.57 | 160.1 | 340.9 |
|  | PA | 7,199 | 2,175 | 0.30 | 105.3 | 123.9 |
|  | AP | 925 | 256 | 0.28 | 137.4 | 145.9 |
|  | TO | 1,236 | 444 | 0.36 | 98.8 | 138.1 |
| Northeast | MA | 4,500 | 983 | 0.22 | 80.9 | 68.7 |
|  | PI | 2,840 | 807 | 0.28 | 110.4 | 122.7 |
|  | CE | 9,993 | 2,281 | 0.23 | 137.3 | 123.4 |
|  | RN | 3,007 | 912 | 0.30 | 105.9 | 128.1 |
|  | PB | 3,672 | 1,261 | 0.34 | 115.5 | 155.3 |
|  | PE | 9,654 | 1,729 | 0.18 | 125.0 | 89.4 |
|  | AL | 2,489 | 709 | 0.28 | 91.2 | 105.4 |
|  | SE | 2,484 | 639 | 0.26 | 134.3 | 136.6 |
|  | BA | 9,129 | 4,110 | 0.45 | 74.6 | 137.1 |
| Southeast | MG | 11,902 | 8,748 | 0.74 | 68.7 | 204.3 |
|  | ES | 5,081 | 1,639 | 0.32 | 152.0 | 199.5 |
|  | RJ | 25,530 | 8,799 | 0.34 | 178.8 | 251.9 |
|  | SP | 46,717 | 17,406 | 0.37 | 119.2 | 186.6 |
| South | PR | 7,974 | 5,611 | 0.70 | 86.2 | 241.9 |
|  | SC | 5,253 | 3,442 | 0.66 | 90.2 | 234.5 |
|  | RS | 8,872 | 6,085 | 0.69 | 96.0 | 265.3 |
| Central West | MS | 2,347 | 1,267 | 0.54 | 104.7 | 223.1 |
|  | MT | 4,520 | 1,850 | 0.41 | 164.0 | 259.3 |
|  | GO | 6,805 | 2,776 | 0.41 | 119.0 | 192.5 |
|  | DF | 4,259 | 857 | 0.20 | 170.8 | 138.6 |
| Brazil |  | 195,072 | 83,362 | 0.43 | 108.8 | 195.4 |

**Table S11.** Variables included in the correlation calculations (**Fig. 4B**). OVW=Percentage of adults who were overweight in each state capital city in 2019; OBT=Percentage of adults who were obese in each state capital city in 2019; NYT=Percentage of adults with hypertension in each state capital city in 2019; HDI=Human development index in 2017; ALG=Political alignment between the governor and the president, as indicated by the governor’s support during the 2018 presidential elections; HId=Median Locational Hoover Index of COVID-19 deaths in 2020; and MRt=Maximum effective reproductive number in 2020 (MRt).

| Region | State | OVW | OBT | HYT | HDI | ALG | HId | MRt |
| --- | --- | --- | --- | --- | --- | --- | --- | --- |
| North | RO | 56.60 | 19.90 | 19.60 | 0.73 | 1 | 45.72 | 1.70 |
|  | AC | 56.60 | 23.30 | 18.50 | 0.72 | 1 | 38.09 | 1.43 |
|  | AM | 60.90 | 23.40 | 18.40 | 0.73 | 1 | 30.42 | 2.11 |
|  | RR | 54.30 | 21.20 | 20.40 | 0.75 | 1 | 28.64 | 1.66 |
|  | PA | 53.30 | 19.60 | 19.30 | 0.70 | 0 | 49.60 | 1.98 |
|  | AP | 53.30 | 22.90 | 23.30 | 0.74 | 1 | 25.21 | 1.91 |
|  | TO | 49.90 | 15.40 | 17.60 | 0.74 | 0 | 57.37 | 1.69 |
| Northeast | MA | 50.30 | 17.20 | 16.90 | 0.69 | 0 | 62.13 | 1.83 |
|  | PI | 52.70 | 17.60 | 22.40 | 0.70 | 0 | 52.08 | 1.58 |
|  | CE | 55.60 | 19.90 | 21.20 | 0.74 | 0 | 47.88 | 2.33 |
|  | RN | 56.60 | 22.50 | 24.50 | 0.73 | 0 | 48.50 | 1.37 |
|  | PB | 54.70 | 20.40 | 25.60 | 0.72 | 0 | 48.16 | 1.73 |
|  | PE | 59.50 | 21.70 | 28.40 | 0.73 | 0 | 40.90 | 1.86 |
|  | AL | 54.40 | 20.00 | 26.80 | 0.68 | 0 | 45.20 | 1.94 |
|  | SE | 53.60 | 20.60 | 25.10 | 0.70 | 0 | 38.98 | 1.68 |
|  | BA | 51.80 | 18.10 | 25.20 | 0.71 | 0 | 51.97 | 1.84 |
| Southeast | MG | 52.50 | 19.90 | 25.80 | 0.79 | 1 | 53.87 | 2.11 |
|  | ES | 49.10 | 17.60 | 24.30 | 0.77 | 0 | 32.81 | 1.50 |
|  | RJ | 57.10 | 21.70 | 28.00 | 0.80 | 1 | 26.20 | 2.57 |
|  | SP | 55.80 | 19.90 | 24.40 | 0.83 | 1 | 31.15 | 2.66 |
| South | PR | 53.70 | 19.40 | 21.10 | 0.79 | 1 | 47.40 | 2.36 |
|  | SC | 53.60 | 17.80 | 21.60 | 0.81 | 1 | 55.21 | 1.81 |
|  | RS | 59.20 | 21.60 | 28.20 | 0.79 | 1 | 48.39 | 1.96 |
| Central West | MS | 58.00 | 22.50 | 24.90 | 0.77 | 1 | 41.97 | 1.61 |
|  | MT | 55.80 | 22.50 | 22.80 | 0.77 | 1 | 51.41 | 1.52 |
|  | GO | 52.70 | 19.50 | 24.30 | 0.77 | 1 | 38.66 | 1.39 |
|  | DF | 55.00 | 19.60 | 28.50 | 0.85 | 1 | 43.23* | 1.47 |

\* Since the Federal District is not divided into municipalities (as other states), we could not calculate the location Hoover Index, and thus we used the median HId for Brazil (13).

